# Distinct adaptations of endocrine and cognitive functions may contribute to high variability in long-term weight loss outcome after bariatric surgery

**DOI:** 10.1101/2022.12.06.22283109

**Authors:** Mathis Lammert, Evelyn Medawar, Hendrik Hartmann, Linda Grasser, Arne Dietrich, Wiebke Fenske, Annette Horstmann

## Abstract

**Background:** Bariatric surgery has been widely recognized as the most efficient long-term treatment method in severe obesity, yet therapy success shows considerable interindividual variability. Postoperative metabolic adaptations, including improved gut hormone secretion (GLP-1, PYY and ghrelin), and restored executive function may play an explanatory role in weight loss, yet causes for poor success in individual patients remain unknown. This study investigates gut-hormonal and cognitive characteristics in extreme weight loss responders to bariatric surgery.

**Methods:** Patients (n=47) with high or low excessive weight loss (EWL) at least 2 years after Roux-en-Y-gastric bypass or sleeve gastrectomy were allocated into good responders (GR, EWL 82.4 ± 11.6%) and poor responders (PR, EWL 24.0 ± *SD* 12.8%) to study differences in postprandial secretion of GLP-1, PYY, ghrelin and in working memory (WM).

**Results:** Mean BMI was 47.1 ± 6.2 kg/m^2^ in poor responders (n=21) and 28.9 ± 3.1 kg/m^2^ in good responders (n=26, *p* < 0.001). Fasted GLP-1 and PYY were comparable for GR and PR (*p* > 0.2) and increased strongly after a standardized test meal (300 kcal liquid meal) with a peak at 15 to 30 minutes. The increase was stronger in GR compared to PR (GLP-1, PYY: *Time* x *Group p* < 0.05). Plasma ghrelin levels already differed between groups at fasted state, showing significantly higher levels for GR (*p* < 0.05). Postprandially, ghrelin secretion was suppressed in both groups, but suppression was higher in GR (*Time* x *Group p* < 0.05). GR showed significantly higher WM scores than PR (*p* < 0.05). Postprandial ghrelin (iAUC), but not GLP-1 or PYY release, significantly mediated the relationship between EWL and a WM subscore (IS score, CI = 0.07 - 1.68), but not WM main score (MIS score, CI = −0.07 - 1.54).

**Conclusion:** Excess weight loss success after bariatric surgical procedures is associated with distinct profiles of gut-hormones at fasted and postprandial state, and differences in working memory. Working memory performance was partly mediated by postprandial incremental drops in ghrelin. Future studies need to integrate longitudinal data, larger samples and more sensitive cognitive tests.

**Highlights:**

- Fasted and postprandial gut hormone release differs between good and poor surgery responders
- Good responders show higher working memory performance
- Postprandial ghrelin dynamics mediate the relationship between excessive weight loss and a subscore of working memory performance
- Longitudinal data are needed to investigate the gut-brain interactions with regard to cognitive functions after bariatric surgery

## 1. Introduction

### Bariatric surgery does not always lead to long-term weight loss

Bariatric surgical procedures remain the most efficient long-term strategy for adiposity reduction in morbid obesity [1–4]. The two most commonly used surgical procedures, Roux-en-Y gastric bypass (RYGB) and laparoscopic sleeve gastrectomy (SG), lead to sustained weight loss, paralleled by decreased morbidity and mortality [4,5]. However, the long-term success of bariatric surgery can vary remarkably between patients. Between 20-30% of patients – so called non-responders – exhibit only minor effects on adiposity reduction or even regain body weight 1-2 years after surgery [6–8]. Mechanisms and/or predictors that determine long-term weight loss success of bariatric surgery remain largely unknown. Understanding such mechanisms would help reduce the burden of 8 invasive therapy in potentially unsuccessful candidates for weight loss due to bariatric surgery and may help in identifying promising molecular targets for novel non-invasive weight loss therapies.

### Changes in hormone secretion might be one mechanism behind weight loss

Mechanistically, weight loss after bariatric surgery is induced by reduced caloric intake, yet cannot be explained solely by food intake restriction (see Reviews by [9,10]). Only weeks after bariatric surgery and before weight loss becomes apparent, alterations in metabolism and eating behavior manifest, i.e., severe improvements in glycemic control [1,2,11], a reduction of hunger sensation, and an increase in post-meal satiety [12], as well as alterations in food preferences [13–15]. In contrast, weight loss due to conventional diets with reduced caloric intake complicates sustained success by increased hunger and decreased satiety [16]. This argues for fundamental effects of bariatric surgery on metabolism and eating behavior which precede and might induce weight loss [17]. The underlying mechanisms of these phenomena have been subject to a great body of research. It appears that the reconfiguration of the gut affects nervous afferents and especially hormonal signals from the intestine to the brain which modify energy homeostasis and eating behavior. In the physiological state, food intake triggers release of hormones such as glucagon-like-1 (GLP-1) and peptide tyrosine tyrosine (PYY) from intestinal cells or inhibits release of stomach-derived ghrelin. These gut hormones signal postprandial metabolic adaptation to the newly ingested energy, including corresponding changes in feelings of hunger and satiety. They act in concert with signals of long-term energy availability such as leptin by exerting effects on receptors in brain regions involved in homeostatic and hedonic eating control. After bariatric surgery, these gut hormones show strongly altered secretion profiles [16,18]. Thus, it is likely that they play a major role in altered gut-brain-communication in individuals after bariatric surgery. For Reviews see [19,20].

### Cognitive impairments associated with obesity might be reversable by bariatric surgery

In the context of obesity, it is striking that individuals show difficulties in implementing planned, goal-directed behavior, e.g., healthier diet and more exercise, in the long-term. The ability to control behavior to adequately achieve a goal relies on a set of central cognitive functions and processes, called executive functions [21]. Central executive functions include a wide palette of abilities, including working memory, response inhibition, set-shifting, reasoning, problem-solving, organizing or planning [22]. Disruptions in nearly all these domains have been shown in individuals with overweight or obesity in a recent meta-analysis [23] and may characterize a neurocognitive profile specific to obesity [21]. One cognitive process responsible for controlling obesogenic behavior may be working memory – a basic construct necessary for short-term storage and manipulation of information until a task is complete. Deficits in working memory in obesity have been reported consistently by a number of studies [24–26]. Interestingly however, they seem to improve after bariatric surgery: Two recent reviews report an overall improvement in executive functioning after bariatric surgery, which was most prominent in memory and working memory [27,28]. Alosco and colleagues [29] used a standardized set of cognitive tasks in 50 candidates of bariatric surgery. At 12 weeks and up until 24 months after surgery, patients showed significant improvements in functions of memory, attention, and executive functions. Although a further follow-up at 36 months showed a decline in some cognitive functions (attention), executive functioning remained significantly improved, indicating sustained postoperative adaptation. The reasons for the postoperatively improved executive functions remain largely unaddressed. However, executive functions and brain areas implicated in control over food intake rely on common neural pathways, which are heavily modulated by neuroendocrine gut hormones [30]. Although these neurobiological links may provide an explanatory model for how the postoperatively altered gut hormone response and executive function contribute to the high inter-individual variability in weight loss after bariatric surgery, there is limited evidence to support this hypothesis.

### A possible mechanism behind variability in weight loss after surgery: Linking gut hormones to the brain

Current evidence suggests that adaptation of gut hormonal release [9,12] and executive function [28] are associated with weight loss after bariatric surgery, yet their impact on treatment outcome variability remains unknown. Cross-sectional models might help to determine whether failure to achieve weight loss associates with blunted adaptation of these factors, when comparing patients with good (GR) and poor therapeutic weight loss response (PR) to bariatric surgery. Although, people in industrialized countries spend a significant portion of their lives in the non-fasted state [31], only few studies examined the dynamics of gut hormone release in response to food intake in GR vs. PR of bariatric surgery [32–35]. Overall, these studies could demonstrate a more pronounced anorectic secretion profile in GR (e.g., stronger release of GLP-1 and PYY and lower release of ghrelin), compared to PR, which may emphasize the role of these hormones in facilitating postoperative weight loss. Yet it remains unclear, if these hormonal adaptions are linked to changes in executive functioning. One study demonstrated reduced inhibitory control in two neurocognitive tests in PR compared to GR [36], but this relationship has not yet been addressed for working memory. To our knowledge, the influence of postoperatively altered gut hormone release on executive functioning in the context of successful and unsuccessful bariatric surgery has not yet been investigated.

### Aims of the study

This study investigates the link between gut hormones and working memory in relation to long-term treatment outcome after bariatric surgery. Comparing patients with high or low excessive weight loss at least two years after bariatric surgery allowed to investigate three questions: (1) Are GR compared to PR in our sample showing a more pronounced postprandial gut hormone secretion? (2) Are there differences in cognitive function between GR and PR? (3) Do gut hormones mediate possible cognitive differences between GR and PR?

## 2. Materials and Methods (sufficient detail for other investigators to repeat the work, appropriate resource deposits)

### 2.1 Participants

Participants were identified and contacted from the bariatric surgery database at the Integrated Research and Treatment Center (IFB) AdiposityDiseases, University Hospital Leipzig, Germany, after screening for the 5% best and worst weight loss responders categorized by excessive weight loss (EWL) at a minimum time span of two years after bariatric surgery at the University Clinic Leipzig, Germany. Included surgical procedures were either laparoscopic Roux-en-Y gastric bypass (RYGB) or laparoscopic sleeve gastrectomy (SG) (for details see **Repository**). Pre-surgery clinical data were collected from the IFB Adiposity database and relevant medical reports. Patients were regularly medically screened after surgery.

Based on information provided in the bariatric surgery database, we grouped patients into GR and PR according to EWL as treatment outcome. EWL was defined as the loss of body-mass-index (BMI) points above normal-weight (obesity defined as BMI > 25 kg/m^2^ [37]) from pre-surgery (2-4 weeks pre-surgery before onset of a specialized preoperative diet) to the last known follow-up measurement as provided by the database and was calculated using (1):

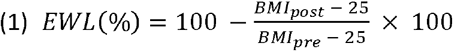

Relative EWL thresholds were initially pre-defined according to previous literature [38] and adapted to fit recruitment requirements with <40% for poor and >60% for good responders of bariatric surgery **(Figure 1A)**.

**Figure 1:**
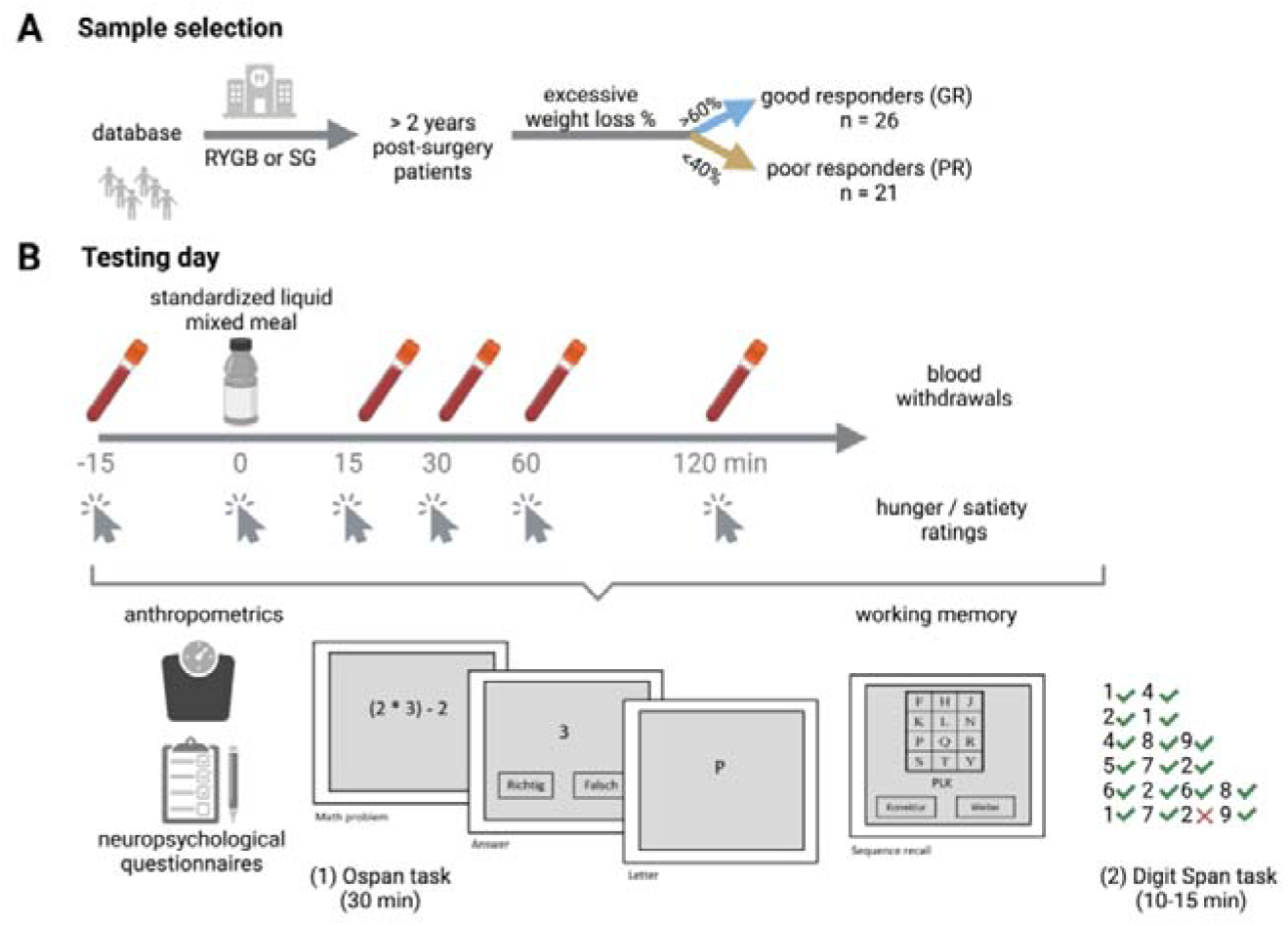
Overview of the study rationale and study design. A) Sample selection overview. B) Overview of testing day for each participant. On the test day, we measured weight and height to calculate BMI and EWL (see formula 1). The illustration for the Automated Operation Span Task was adapted from Unsworth et al [40]. This figure has been created with BioRender.com. Abbreviations as follows: RYGB: Roux-en-Y gastric bypass, SG: sleeve gastrectomy.

Exclusion criteria for this study were acute neurological and/or psychiatric diseases, intake of dopaminergic medication, history of neurosurgical interventions or severe head trauma, abuse of alcohol and/or drugs, pregnancy/lactation or insufficiently adjusted diabetes mellitus according to medical history. Out of 53 initially recruited patients, 6 participants were excluded in retrospect for the following reasons: History of additional bariatric surgery (n=2), history of cerebral trauma (n=2), early test abortion due to medical issues (n=1) or history of long-time corticosteroid intake (n=1). Two PR datasets were incomplete due to missing postprandial gut hormonal data because of a strong aversion towards the test meal (n=1) and early test abortion due to personal issues (n=1). Both datasets were excluded from the respective analyses (flow chart see **Supplementary Figure 1**).

### 2.2 Study design

The study design has been described previously [39]. Participants were invited for a single testing day at the Max Planck Institute for Human Cognitive and Brain Sciences (MPI CBS), Leipzig, Germany. **Figure 1B** describes all procedures of the test day. In brief, patients provided blood samples in fasted state (overnight fasted, minimum 10 hours), after which they consumed a standardized liquid mixed meal (300 kcal, 125 ml). Additional blood samples were taken 15, 30, 60, and 120 minutes after ingestion of the standardized meal to measure the postprandial response of gut hormones. At each time point, hunger and satiety were self-reported on a digital visual analog scale. During these 2 hours and after that, participants completed self-reported questionnaires and cognitive measures of working memory (Operations span (Ospan) task and Digit span task) in a standardized sequence across participants.

### 2.3 Serum markers and subjective hunger/satiety ratings after a standardized mixed meal

Participants were fasted overnight (10 hours minimum) and blood samples were taken immediately before (0 min) and 15, 30, 60 and 120 min after a standardized liquid mixed meal (125 ml, 300 kcal containing 12 g of protein, 12 g of fat and 37 g of carbohydrates, Nutricia Fortimel Compact, Nutricia Milupa GmbH, Hamburg, Germany).

For gut hormone analysis, tubes were prefilled with aprotinin (500 KIU per ml blood) to avoid enzymatic hormone degradation. Tubes were cooled at 4°C until centrifugation at time points 60 and 120 min. After centrifugation at 4°C for 10 min at a speed of 3000 rpm, plasma was pipetted to 2ml Eppendorf tubes and stored at −80°C. Measurements were based on plasma and performed by LaboPart (Laborservice GmbH, Elsterwerda, Germany), including total GLP-1, total PYY, insulin and leptin, that were performed simultaneously in a commercial multiplexing kit (Milliplex MAP Human Metabolic Hormone Magnetic Bead Panel, Merck, CAT # HMHEMAG-34K). Total ghrelin was measured using Human Ghrelin (Total) ELISA Kit (Merck, CAT # EZGRT-89K). Each measurement was performed in duplicates, and mean results were used. Values were curated for lower and upper detection limits and several data points were missing due to unsuccessful blood drawing or invalid measurements (see **Repository**). Prior to each blood withdrawal, participants were instructed to rate their feelings of hunger and satiety based on digital visual analog scales (range 0 – 10; hunger: 0 – not hungry, 10 – very hungry; satiety: 0 – not sated, 10 – very sated) as well as individual palatability of the meal (0 – not palatable; 10 – very palatable).

Additional overnight fasted blood samples were taken from serum and EDTA tubes to measure glycated hemoglobin (HbA1c), glucose, insulin, triglycerides (TG), cholesterol, high- and low-density lipoproteins (HDL and LDL, respectively), aspartate transaminase (ASAT), alanine transaminase (ALAT), C-reactive protein (CRP) and interleukin 6 (IL-6). Based on fasting glucose and fasting insulin we calculated insulin resistance with the homeostasis model assessment (HOMA-IR [41]) using the formula: HOMA-IR = fasting glucose [mmol/l] × fasting insulin [pmol/l] / 135.

Further, as part of the preoperative standard evaluation, the following parameters had been assessed before surgery: Hb1Ac, glucose, insulin, LDL, HDL, TG, cholesterol, ASAT, ALAT and CRP. These blood samples were analyzed at the Institute for Laboratory Medicine, Clinical Chemistry and Molecular Diagnostics, University Hospital Leipzig, Leipzig, Germany. Note, that for pre-operative timepoint we encountered numerous missing values (see **Supplementary Table 6**).

### 2.4 Executive function

We assessed working memory as a component of executive function in two tasks.

The Digit Span Task (DST) is part of the Wechsler Adult Intelligence Scale [42] and represents a simple measure of working memory and attention. Participants had to recall sequences of digits that were displayed via audio playback from a computerized version of the task (10 to 15 min). Sequence lengths increased every second length. The test ended if one length was not recalled correctly twice. The final score was the longest recalled sequence. We used DST forward and DST backward (recall in inversed order).

We used a modified version of the automated operation span (OSPAN) task that has been used in our lab and was described in detail before [40,43]. During the OSPAN task (30 min), participants were instructed to memorize sequences of letters, ranging from 3 to 7 items, while intermittently being distracted by math problems. Before the screening of each letter for 800 ms, a simple math operation was presented followed by a two-alternative forced choice to report the correct solution. At the end of each trial, participants were asked to recall the letters in correct order on the screen by mouse click. The presentation of sequences was randomized. Each sequence length was presented three times, summing up to a total of 15 sequences, including 75 letters and 75 math problems. The test trials were preceded by 3 practice trials to separately rehearse execution of letter and math tasks, as well as and the combination of letter- and math task. Within the math practice trial, the mean time for solving the math problems was computed for each patient, which was then used to define the maximum time allowed for submitting the solution of math problems in the actual task (maximum time = mean time + 2.5 SD). Working memory capacity was computed using the Math-Item-Sequence (MIS) and the Item-Sequence score (IS) [44]. The MIS score captures possible trade-offs between actual working memory and increased focus on the mathematical problems; the IS score assesses only the working memory components of the OSPAN task. The minimum performance is zero and the maximum performance is 15 for both MIS and IS.

### 2.5 Self-reported questionnaires

Questionnaires assessed self-reported depressive symptoms (BDI-II), psychosocial traits like eating behavior (EDE-Q, DEB-Q, DFS-Q), personality traits (NEO-FFI), chronic stress (TICS) and emotional regulation (DERS), history of smoking (FTND), and neurocognitive domains like impulsivity (UPPS), delayed gratification (MCQ), and approach/avoidance behavior (BIS/BAS) (details and data in **Repository**).

### 2.6 Statistical analyses

We used R version 3.4.3 within RStudio [45] for all statistical analyses. The significance level alpha was 0.05 for all statistical tests.

For testing mean differences of groups (weight status, demographics, questionnaire scores), we either used Student’s t-test for independent samples or Whitney-Mann-U test, if assumptions of normality and homogeneity of variance were not met. Normality was tested with the Shapiro-Wilk test and homogeneity of variance with Levene’s test. For group distributions of categorical variables (sex, type of surgical procedure), Pearson’s Chi-square test was employed. Outliers were defined as values below or above a 2.2 interquartile range from the whole sample’s lower or upper quartile, respectively [46] and in few cases excluded after individual evaluation, as stated explicitly.

For repeated-measure designs, we used linear mixed effects models (LME) to model best fit by maximum likelihood estimation (lme function in ‘nlme’ package [47]). LME modelling was suggested for the field of bariatric surgery [48] as a more accurate approach in repeated-measures designs and we used it in three occasions: (1) To assess group differences of metabolic blood markers before and after surgery. The model included plasma level (e.g., HbA1c, defined as outcome), time point of measurement (main effect *Time*; pre vs. post) and weight-loss group (main effect *Group*; GR vs. PR) as random and fixed effect, respectively, and the interaction term *Time X Group*. (2) To assess group differences in gut hormonal release after food intake. The plasma level was set as outcome, time point of measurement (main effect *time*; 0, 15, 30, 60, 120 min) as random effect and weight loss group (main effect *Group*) and the interaction term *Time X Group* as fixed effects. To reach normal distribution, GLP-1, insulin, and leptin were log-transformed and influential outliers were excluded (insulin n = 1, leptin n = 1) before analysis. (3) To assess group differences in VAS rating of satiety or hunger after food intake. This model was built like the before mentioned analysis of gut hormones.

To further investigate the individual’s dynamic hormone secretion response, we calculated areas under the curve (AUC) over the total measurement period of 120 minutes, using the trapezoidal method [49]. To focus on postprandial dynamics only, additional incremental areas under the curve (iAUC) were computed as AUC over baseline (measure at 0 min) hormone level. Bivariate correlation analyses were performed for iAUC values of hunger ratings and gut hormone levels using Spearman’s rank test and Holm’s adjustment [50] for multiple testing.

For cognitive outcome comparison between weight loss groups, we used either t-test or Mann-Whitney-U test if the assumptions of normality and homogeneity of variance were not met. Outliers based on the 2.2 interquartile range criterion were removed before analysis.

Multiple mediation was used to investigate whether the effect of weight loss on cognitive outcome would be mediated by the gut hormonal response to food intake by using weight loss group as independent variable, postprandial gut hormone secretion patterns of GLP-1, PYY and ghrelin as parallel mediators, and working memory scores as dependent variables. The indirect effect of gut hormonal excretion was computed via bias-corrected bootstrapping method [51,52] and generation of unstandardized results was performed by *sem*-function from ‘lavaan’-package [53] with 5,000 iterations and maximum likelihood estimation of missing data points.

### 2.7 Ethics

The study was accomplished in accordance with the Declaration of Helsinki and was approved by the ethics committee of the University of Leipzig (027/17-ek). Participants gave their written, informed consent before taking part in the study and were recompensed with 9-10 € per hour.

## 3. Results

### 3.1 Group descriptions

We included 47 patients (age: *M* = 51.8 years, *SD* = 10.1 years, 31 to 70 years) that underwent bariatric surgery at least two years before testing (*M* = 4.76 years, *SD* = 1.60 years, 2.11 to 9.64 years, **Figure 2A**). RYGB was performed on 85 % (n = 40 of 47) of the participants (for details on surgical procedures see **Repository**). Female participants accounted for 77 % (n = 36 of 47) of the sample **(Table 1)**. The final sample consisted of 21 poor responders (EWL=24.0%, *SD*=12.8%) and 26 good responders (EWL=82.4%, *SD*=11.6%). Mean BMI at test date was 47.1 ± 6.2 kg/m^2^ in PR and 28.9 ± 3.1 kg/m^2^ in GR (*p* < 0.001, see **Figure 2B**). There were no significant group differences in age, time since surgery, education years, proportions of gender or type of surgical procedure **(Table 1)**. For information on medical history, we screened comorbidities, medication intake (most commonly antihypertensives, antidiabetic medication or proton pump inhibitors) and medical status (reported in detail in **Repository**).

**Table 1:** Descriptive statistics for good and poor weight loss responders of bariatric surgery.

|  | Poor Responder (n=21) |  |  | Good Responder (n=26) |  |  | Test statistics |  |
| --- | --- | --- | --- | --- | --- | --- | --- | --- |
|  | *n* | % |  | *N* | % |  | *Statistic* | *p* |
| Female sex | 16 | 76.2 |  | 20 | 76.9 |  | 0.00<sup>c</sup> | 0.953 |
| RYGB | 17 | 81.0 |  | 23 | 88.5 |  | 0.52<sup>c</sup> | 0.472 |
|  | *Mean* | *SD* | *Range* | *Mean* | *SD* | *Range* | *Statistic* | *p* |
| Age (years) | 52.9 | 10.5 | 31 - 67 | 50.9 | 9.8 | 33 - 70 | 313<sup>b</sup> | 0.403 |
| Education (years) | 13.5 | 1.6 | 12 - 17 | 13.7 | 1.86 | 10 - 17 | 229<sup>b</sup> | 0.280 |
| Years after surgery | 4.7 | 1.4 | 2.1 - 6.5 | 4.8 | 1.76 | 2.1 - 9.6 | -0.07<sup>a</sup> | 0.941 |
| Preoperative BMI (kg/m<sup>2</sup>) | 54.6 | 8.4 | 41.6 - 74.6 | 46.7 | 7.41 | 34.0 - 63.7 | 3.43<sup>a</sup> | **0.001** |
| BMI on test day (kg/m<sup>2</sup>) | 47.1 | 6.2 | 40.3 - 61.9 | 28.9 | 3.13 | 25.1 - 37.5 | 546<sup>b</sup> | **< 0.001** |
| Weight loss (%) | 13.1 | 7.5 | 1.0 - 25.7 | 37.1 | 8.38 | 16.5 - 54.2 | -10.2<sup>a</sup> | **< 0.001** |
| Weight loss (kg/m<sup>2</sup>) | 7.5 | 5.0 | 0.4 - 19.1 | 17.8 | 6.17 | 5.6 - 33.1 | -6.19<sup>a</sup> | **< 0.001** |
| EWL (%) | 24.0 | 12.8 | 2.6 - 40.0 | 82.4 | 11.6 | 62.2 - 99.5 | -16.4<sup>a</sup> | **< 0.001** |
| HbA1c (%) | 5.97 | 0.92 | 5.0 - 8.0 | 5.14 | 0.65 | 4.4 - 7.8 | 465<sup>b</sup> | **< 0.001** |
| Fasting glucose (mmol/l) | 6.52 | 1.66 | 4.8 - 11.5 | 5.17 | 0.56 | 4.3 - 6.8 | 443<sup>b</sup> | **< 0.001** |
| Insulin (pmol/l) | 82.1 | 45.0 | 26 - 234 | 46.7 | 32.7 | 11 - 167 | 442<sup>b</sup> | **< 0.001** |
| HOMA-IR | 4.02 | 2.89 | 1.0 - 15.0 | 1.82 | 1.45 | 0.6 - 7.3 | 471<sup>b</sup> | **< 0.001** |
| LDL (mmol/l) | 2.49 | 0.47 | 1.7 - 3.3 | 2.37 | 0.56 | 1.6 - 4.0 | 327<sup>b</sup> | 0.257 |
| HDL (mmol/l) | 1.44 | 0.4 | 0.5 - 2.1 | 1.84 | 0.64 | 1.2 - 3.3 | 192<sup>b</sup> | 0.083 |
| Triglycerides (mmol/l) | 1.40 | 0.4 | 0.7 - 2.2 | 0.89 | 0.39 | 0.5 - 2.1 | 466<sup>b</sup> | **< 0.001** |
| Cholesterol (mmol/l) | 4.18 | 0.64 | 3.0 - 5.2 | 4.31 | 0.9 | 2.9 - 6.2 | -0.55<sup>a</sup> | 0.584 |
| ALAT (μkat/l) | 0.37 | 0.15 | 0.2 - 0.7 | 0.41 | 0.18 | 0.1 - 1 | 244<sup>b</sup> | 0.542 |
| ASAT (μkat/l) | 0.40 | 0.11 | 0.2 - 0.7 | 0.5 | 0.22 | 0.3 - 1.5 | 183<sup>b</sup> | 0.054 |
| CRP (mg/l) | 12.8 | 32.7 | 0.7 - 153 | 0.65 | 0.75 | 0.0 - 3.6 | 509<sup>b</sup> | **< 0.001** |
| IL-6 (pg/ml) (n=21:25) | 6.84 | 9.97 | 1.3 - 49.3 | 2.17 | 2.96 | 1.3 - 15.5 | 469<sup>b</sup> | **< 0.001** |

Note: Proportions of gender and type of surgical procedure are reported in number and percent. Numbers of compared datasets in brackets (PR:GR), if incomplete. Significant values $p < 0.05$ are printed in bold. <sup>a</sup> Independent t-test, <sup>b</sup> Mann-Whitney-U test, <sup>c</sup> Pearson's Chi<sup>2</sup>-test. BMI body-mass-index, EWL excessive weight loss.

**Figure 2:**
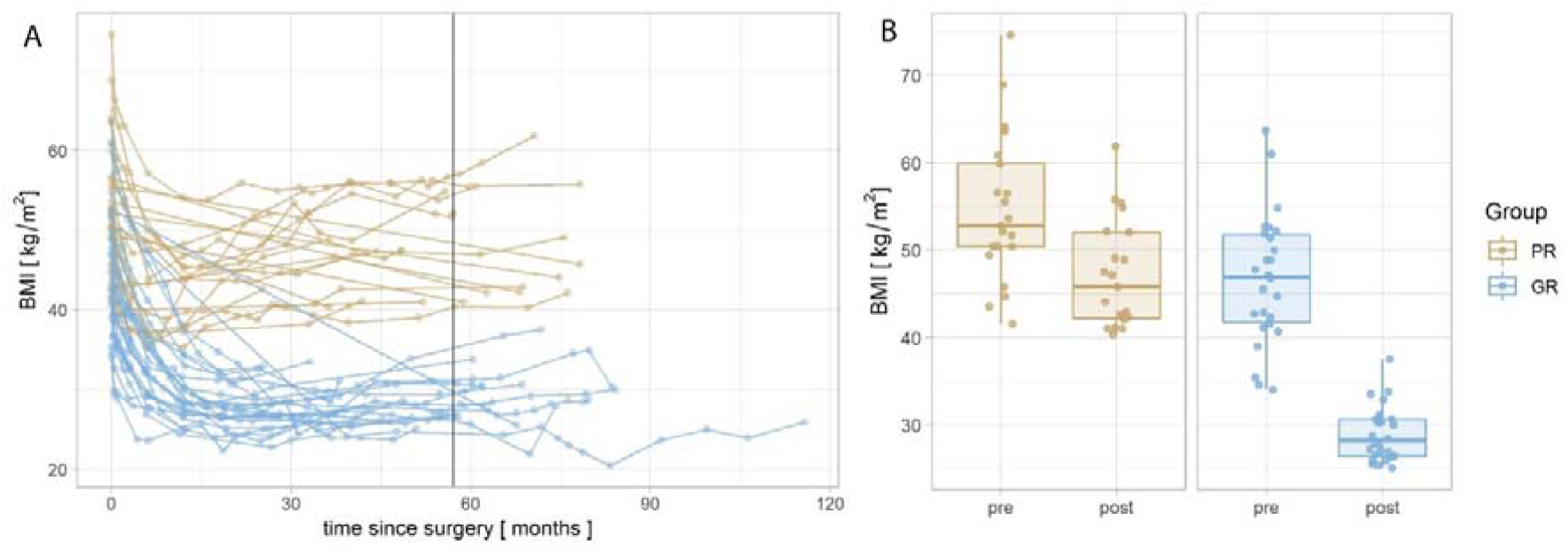
A) BMI trajectory from time point of bariatric surgery until test date. Points connected by lines indicate follow up measurements of one individual. The black vertical line indicates sample mean time since surgery at test date. B) BMI before bariatric surgery (“pre”) and at test date (“post”). Points indicate individual data points; boxplots indicate statistical dispersion. A/B) Color codes for weight loss groups: Poor Responders (PR, brown) and Good Responders (GR, blue).

### 3.2 Postprandial gut hormonal release of both weight loss groups

Postprandial release of GLP-1 and PYY into systemic circulation increased significantly (main effect time, GLP1: *X*^2^ = 239, *p* < 0.001; PYY: *X*^2^ = 129, *p* < 0.001) in both study groups, however, this release was significantly higher in good responders (interaction effect *group x time*, GLP1: *X*^2^ = 10.7, *p* < 0.05; PYY: *X*^2^ = 11.1, *p* < 0.05, see **Figure 3**). Ghrelin release after standardized test meal was significantly suppressed in both groups (main effect *time X*^2^ = 115, *p* < 0.001), but stronger in good responders (interaction effect *group x time X*^2^ = 17.7, *p* < 0.001). Notably, at baseline, good responders showed higher levels of ghrelin (*U* = 139, *p* < 0.05), but not of GLP-1 and PYY (*p* > 0.05), compared to poor responders. Additionally, postprandial release of insulin showed a significant increase with peak levels after 30 minutes (main effect *time X*^2^ = 236, *p* < 0.001), but did not differ between groups (interaction effect *group x time X*^2^ = 5.39, *p* > 0.05) or at baseline (*U* = 352, *p* = 0.050). Leptin levels of good responders were significantly lower at baseline (*U* = 507, *p* < 0.01). While there was a significant change in leptin plasma levels over time (main effect *time X*^2^=17.8, *p*<0.001), this effect did not differ between groups (interaction effect *group x time X*^2^ = 6.09, *p* > 0.05; **Figure 3** below; baseline-corrected see **Supplementary Figure 2** and **Supplementary Table 1** for descriptives of blood markers of each postprandial time point in **Repository**).

**Figure 3:**
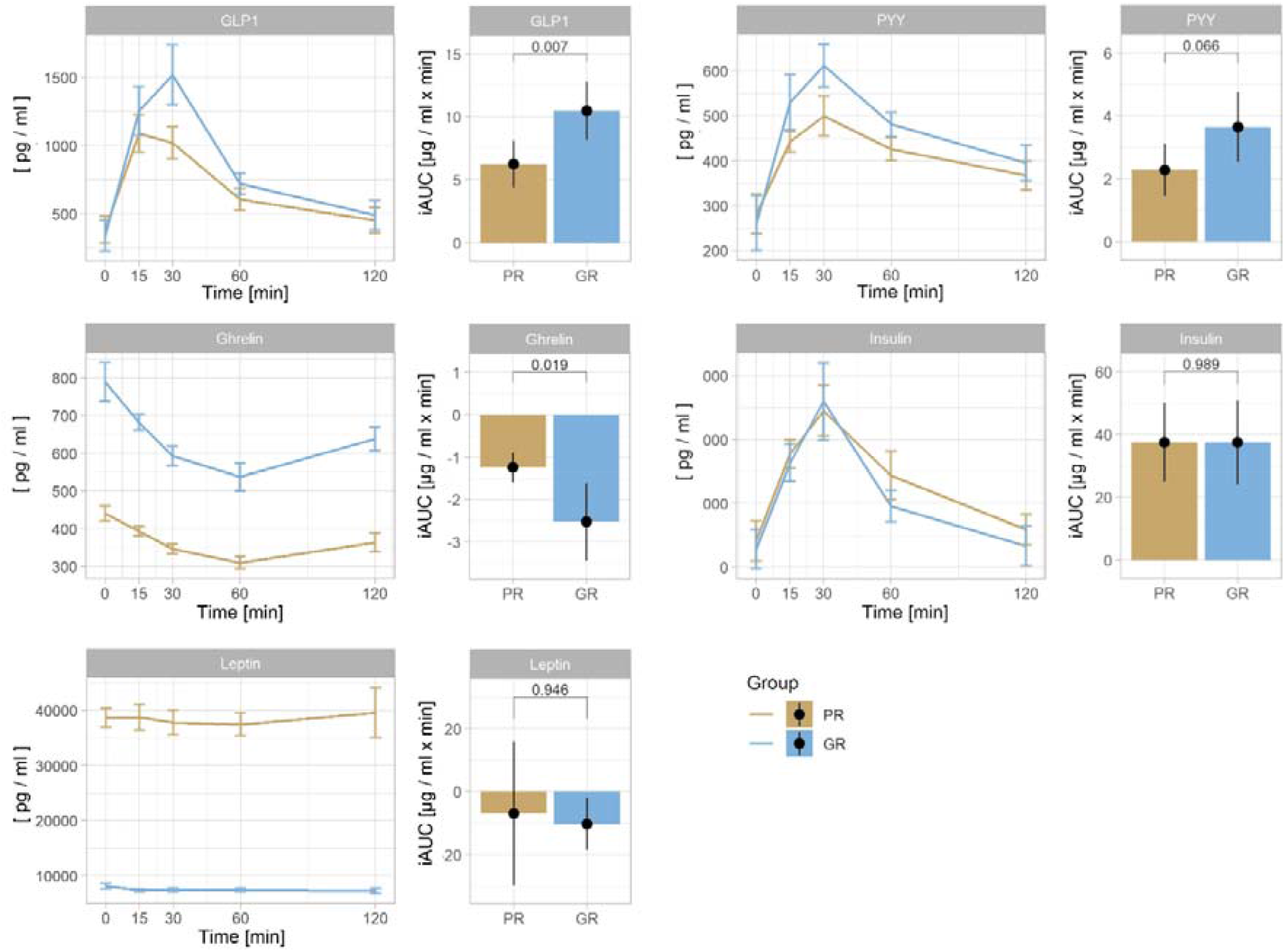
Postprandial gut hormone plasma levels depicted by weight loss group. Line plots (left) show absolute plasma levels of glucagon-like-peptide 1 (GLP-1), peptide YY (PYY), ghrelin, insulin, and leptin before (0 min) and 15, 30, 60 and 120 min after food intake for good (GR) and poor weight loss responders (PR) of bariatric surgery; error bars indicate within-subject 95% confidence interval. Box plots (right) show incremental area under the curve (iAUC; relative to fasted plasma levels) in [μg / ml x min] of gut hormones for good and poor weight loss responders; the number reflects p-value of group comparisons. Demonstrated data includes outliers.

In addition, results for iAUC (iAUC = AUC minus levels at fasted state) overall replicated the LME model findings presented above, with significant group differences for GLP-1, ghrelin and a trend for PYY, and are depicted in **Figure 3** (Details in **Supplementary Table 2** in the **Repository**).

### 3.3 Subjective hunger and satiety rating and correlation to gut hormone release of both weight loss groups

LME models revealed a significant postprandial increase in satiety (main effect *time X*^2^ = 51.8, *p* < 0.001) and a decrease in hunger ratings (main effect *time X*^2^ = 25.4, *p* < 0.001) in both weight loss groups, which did not differ between groups (interaction effect *group x time*, hunger: *X*^2^ = 3.41, *p* > 0.05, satiety *X*^2^ = 4.9, *p* > 0.05). At fasted state, there was no significant group difference in both ratings (hunger *U* = 193, *p* > 0.05, satiety *t*(45) = 0.89, *p* > 0.5), but a trend towards more hunger at all time points and earlier postprandial loss of satiety after 2 hours in good responders (see **Supplementary Figure 3** and **Supplementary Table 3** in the **Repository**). Pleasantness of meal was rated equally across groups (GR 3.86 ± 2.75, PR 5.1 ± −3.19, *U* = 210, *p* > 0.05).

Correlation analysis of baseline-corrected iAUC postprandial VAS ratings and gut hormone releases revealed a significant negative correlation for hunger ratings with GLP-1 (*r*_*s*_ = −0.36, *p* < 0.05) and ghrelin release (*r*_*s*_ = 0.33, *p* < 0.05), but not with PYY, insulin or leptin. After controlling for multiple comparisons, no correlation remained significant. Satiety ratings did not significantly correlate with any gut hormone release (**Supplementary Table 4** in **Repository**).

### 3.4 Working memory capacity in parts higher in good compared to poor weight loss responders

Good responders showed a significantly higher MIS score than PR to bariatric surgery, *t*(43) = −2,04, *p* = 0.048 (see **Figure 4**). The IS score closely failed statistical significance in a non-parametric test, *U* = 168, *p* = 0.062. The Digit Span score did not show a significant group difference in both sub scores, therefore it was disregarded for consecutive mediation analyses (forward subscore: *U* = 215, *p* = 0.302; backward subscore: *U* = 229, *p* = 0.469).

**Figure 4:**
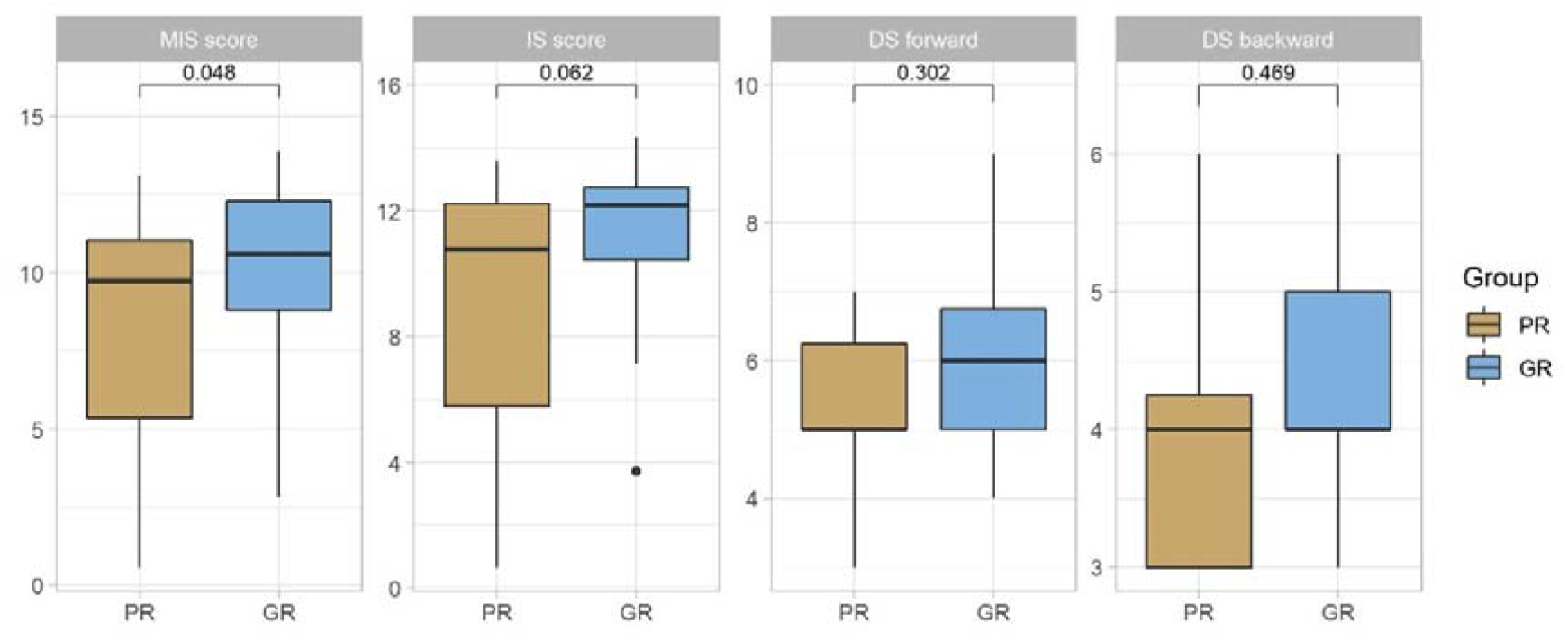
Working memory scores between weight loss groups as generated by Ospan (MIS and IS score) and Digit Span task (DS; forward and backward score) including *p*-value. Boxplots show scores, outliers (point) are defined as 2.2 inter-quartile range.

### 3.5 Gut hormones partly mediate the distinct working memory capacity of weight loss groups after bariatric surgery

We investigated a possible mediating effect of gut hormones (accumulated postprandial release (iAUC) of GLP-1, PYY or ghrelin) on weight loss group-dependent differences in working memory scores using two multiple mediation models to account for both measures of working memory (MIS and IS score) **(Figure 5)**. For both scores, weight loss groups showed a significantly positive relationship (path a) with GLP-1 (*β* > 4.10, *p* = 0.004) and ghrelin (*β* = −1.3, *p* < 0.01), but not with PYY (*β* = 1.14, *p* > 0.05). That means, good weight loss response was associated with higher postprandial excretion in GLP-1, a trend towards more excretion of PYY and lower excretion of ghrelin. The relationship between PYY response and both MIS scores (path b) was significant (*β* > 0.62, *p* = 0.002), which means that greater postprandial secretion was associated with higher working memory scores. While a higher GLP-1 response was not significantly associated with WM score in neither model, ghrelin showed a more complex relationship. Postprandial ghrelin secretion was significantly associated with IS score (*β* > −0.48, *p* < 0.05), but not with MIS score (*β* = −0.35, *p* = 0.167). The relationship between weight loss success and cognitive outcome (total effect c) was significant in the prediction models of MIS score (*β* > 1.90, *p* < 0.05) and IS score (*β* > 2.16, *p* < 0.05). After accounting for the indirect effects of the suggested mediators, the direct effect between weight loss group and cognitive outcome was no longer significant in none of the models (*p* > 0.5).

**Figure 5:**
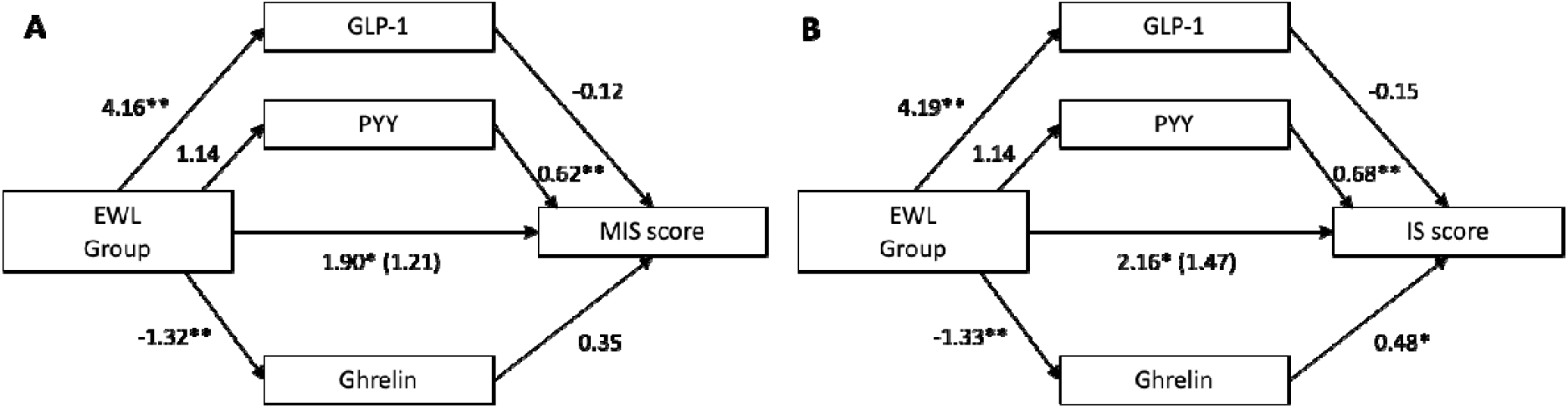
Multiple mediation models investigating the mediating effects of postprandial gut hormone release (postprandial release of GLP-1, PYY and ghrelin as mediators, expressed as iAUC) on the relationship between excessive weight loss group and MIS score (A) and IS score (B). β is reported on each path, including level of significance (* p < 0.05, ** p < 0.01, *** p < 0.001).

The indirect effects for gut peptides’ mediation models are shown in **Table 2**. Ghrelin response significantly mediated the relationship between weight loss group and IS score (estimate = 0.64 (unstandardized = 0.10), SE = 0.38, CI = 0.07-1.68), whereas GLP-1 and PYY response did not. The model using MIS score as outcome showed no significant indirect effect of neither mediator.

**Table 2:** Mediating indirect effects of postprandial gut hormone release (GLP-1, PYY and ghrelin) on the relationship between weight loss group and working memory (MIS and IS score).

| Indirect mediation effects | Estimate | SE | LL 95% CI | UL 95% CI |
| --- | --- | --- | --- | --- |
| **MIS score** |  |  |  |  |
| GLP-1 | -0.49 (-0.08) | 0.56 | -1.94 | 0.36 |
| PYY | 0.71 (0.11) | 0.46 | -0.04 | 1.78 |
| Ghrelin | 0.46 (0.07) | 0.38 | -0.07 | 1.54 |
| **IS score** |  |  |  |  |
| GLP-1 | -0.64 (-0.96) | 0.58 | -2.17 | 0.15 |
| PYY | 0.69 (0.11) | 0.46 | -0.08 | 1.78 |
| Ghrelin | **0.64 (0.10)** | **0.38** | **0.07** | **1.68** |

*Note:* Significant indirect effects (if 95% CI does not cross zero) in bold. Estimate in absolute and standardized (in brackets) values. SE: standard error; LL: lower level; UP, upper level; CI: confidence interval.

### 3.6 Eating behavior and personality traits

We assessed eating behavior and smoking addiction, psychological state and personality traits, which might confound metabolic and cognitive group characteristics (see **Supplementary Table 5** in **Repository**). Ten participants reported smoking habits, but were distributed equally across groups (n = 4 GR / 6 PR, *X*^2^(1) = 0.11, *p* = 0.737). Within these, there was no significant group difference in nicotine dependence, *U* = 21.0, *p* = 0.065. There were moderate depressive symptoms (BDI score = 21 in n = 2) in two participants and no or only mild depressive symptoms in all others without significant group differences, *U* = 346, *p* = 0.117. Poor responders showed significantly higher scores in four eating disorder categories (restraint eating, eating concern, weight concern, shape concern, all EDE-Q subscales *p* < 0.05). While both groups did not differ in the consumption frequency of food containing only saturated fat or only added sugar (HFS, all *p* > 0.2), GR did consume significantly more often products that contained both, high amounts of saturated fat and added sugar, compared to PR (*U* = 390, *p* = 0.013), as assessed by DFS-Q. Emotional eating scores (DEB-Q-EE) were similar across groups (*U* = 276, *p* = 0.966). Personality traits (NEO-FFI) were comparable between groups (all *p* > 0.1) in all domains except for *Extraversion*, where GR showed significantly higher scores (U = 383, *p* = 0.019). There was no group difference in measures of chronic stress (TICS, *U* = 317, *p* = 0.356) or emotion regulation (all DERS subscales, *p* > 0.2). Both weight loss groups did not show differences in measures of delayed gratification of reward (MCQ), impulsivity (UPPS) and approach-avoidance tendencies (BIS/BAS) (all *p* > 0.5).

### 3.7 Distinct metabolic profiles between weight loss groups for pre- and post-surgery

Postoperative parameters of glucose, lipid and hepatic metabolism as well as markers of inflammation were evaluated to characterize both extreme weight loss groups. Poor responders showed significantly higher plasma levels of glucose metabolism (HbA1c and fasted glucose), insulin and insulin resistance (HOMA-IR), compared to GR (p<0.001, see **Table 1**). Lipid metabolism (HDL, LDL and cholesterol) did not differ significantly between both groups, except for higher triglycerides in PR than in GR (*U* = 466, *p* < 0.001). Hepatic transaminases were at physiological levels in both groups (*p* > 0.05). Inflammatory parameters (CRP and IL-6) were significantly elevated in PR, compared to GR (p < 0.001, see **Table 1**).

Preoperative metabolic parameters were available for a subpopulation (n ≥ 15 depending on marker) and allowed for longitudinal analysis across weight loss groups by using LME models (**Supplementary Table 6, Supplementary Table 7 and Supplementary Figure 4** in **Repository**). A significant reduction from pre- to post-surgery was seen in levels of HbA1c, glucose, insulin, HOMA-IR, LDL, triglycerides, cholesterol, ALAT, ASAT and CRP across all participants, while HDL significantly increased (main effect *time* p_all_ < 0.05). Yet, this difference was not significantly larger in GR compared to PR, except for fasting glucose and HDL (interaction effect *group x time, p* < 0.05). Notably, preoperative levels of fasting glucose, insulin and HOMA-IR were significantly elevated in PR (*p*_*all*_ < 0.05).

## 4. Discussion

### 4.1 Short summary of aim and relevance

In a post-hoc design, we aimed to identify differences in cognitive and metabolic outcome between patients with good and poor weight loss success at least two years after bariatric surgery. As hypothesized, we found higher postprandial secretion of GLP-1 and PYY, stronger suppression of ghrelin release and to a certain extent higher cognitive function in good compared to poor responders, which was partially mediated by postprandially suppressed ghrelin levels.

### 4.2 Stronger dynamics of gut peptides and hunger in good responders

According to our hypothesis, we found that the release of GLP-1 and PYY, and the postprandial suppression of ghrelin release was more pronounced in GR, compared to PR of bariatric surgery. Several studies have used similar paradigms, reporting mostly similar postprandial gut-hormone release patterns: Higher GLP-1 release in GR versus PR has been reported most consistently [33,34,54,55] together with a higher release of PYY in most [33,54], but not all studies [34]. The deviating finding in the latter study might be due to less extreme EWL groups, compared to our study (PR EWL mean 24% compared to 35% [34]). Our study did not show differences in fasted levels of GLP-1 and PYY between weight loss groups, which highlights the impact of food intake on those hormones and the importance of including meal-challenges in the study design to map physiological dynamics. For ghrelin, we found higher circulating fasted levels in GR compared to PR. These results are in line with other post-RYGB human studies [34,56–58] and seem to relate to fat mass, although the exact mechanism remains unknown and results differ dependent on surgery type [59]. Following SG, circulating ghrelin levels decrease due to the removal of the ghrelin-producing gastric fundus, but slightly increase in the cause of months postoperatively [60]. Accordingly, a meta-analysis across 53 studies found higher fasted ghrelin levels after RYGB compared to SG mostly after 6 to 12 months post-surgery [61]. In our study, we could not account for surgery type due to the small sample size. Yet, patients receiving SG or RYGB were distributed equally between both weight loss groups in our study and time after surgery was longer in our study, than in most (on average >4.5 years) [59]. The higher postprandial suppression of ghrelin in GR was in accordance with a more pronounced anorexigenic profile of GR, compared to PR, and with other studies on extreme weight loss response (significant higher reduction in GR [33,34], but not in all [55]). The dynamic of postprandial release of insulin was similar to that of GLP-1, which is physiologically expected as GLP-1 triggers insulin secretion under glycemic control. However, we did not see a stronger postprandial release of insulin in GR. This has been reported before [34]. Further, the absence of a group difference in postprandial insulin release might be explained by reduced insulin sensitivity and consecutively increased insulin release in PR, compared to GR. Fasted leptin levels were markedly higher in in PR compared to GR, corresponding to higher levels of fat mass. Physiologically, leptin regulates hunger and eating behavior via central nervous feedback loops [62,63]. These systems seem to be disrupted in obesity with growing neuronal leptin resistance [64], yet might be restored after RYGB [65–67]. Dynamics of postprandial leptin secretion have barely been studied, yet a dysregulation in obese versus normal-weight individuals has been shown [31]. Our data could not show differences by bariatric surgery success for postprandial leptin secretion.

The subjective perception of hunger and satiety, assessed by VAS scales at each blood collection time point, paralleled the expected effect after food intake (decrease in hunger and increase in satiety) in both groups. GR tended to report higher levels of hunger and earlier decreases in satiety. However, possibly due to the high variability of the VAS ratings ranging from 0 to 10 in both groups and at all time points, no statistical significance was reached. The wide range of data must be interpreted carefully, partly possibly due to erroneous data entry by the participants, but could also be related to individual paradoxical feelings of satiety and hunger after bariatric surgery, which has been reported anecdotally by participants of this study and systematically by other studies [10,12]. After food intake, feedback loops of gut hormones such as GLP-1, PYY and ghrelin lead to a reduction of hunger and an increase of satiety [68]. We could demonstrate this association between hunger with ghrelin and GLP-1, but not with PYY, insulin or leptin, and all associations became insignificant after controlling for multiple comparison. As a reason for this, in addition to the above-mentioned high variability in VAS ratings, it can be assumed, that correlating a single gut peptide with hunger or appetite does not reflect the complex and often synergistic effects of hormones and other signals, which regulate eating behavior [10].

Taken together, we found that anorexigenic gut hormone profiles were more pronounced in GR than in PR, adding correlational evidence to the hypothesis of a mechanistic link between postoperative gut-hormonal adaptations and weight loss.

### 4.3 Weight loss groups differ in aspects of working memory function

As hypothesized, we found significantly higher working memory capacity in the good responder group compared to the bad responder group. This is in line with proposed differences in cognitive function relating to weight status [21]. The effect was most prominent in overall MIS score of the Ospan task but absent in subscores of the Ospan or Digit span task. The Ospan task, including MIS score [43,44], is a robust and valid tool for measuring working memory capacity. Its IS subscore can be regarded as a more direct measure of the Ospan task’s working memory capacity component compared to MIS score, which includes performance of the distractor task. It remains unclear whether the non-significant result for IS score is due to the more conservative, yet more robust, non-parametric testing or the difference in complexity. Even though there were no group differences of known confounding factors of executive function i.e., age, education and depressive symptoms, our sample includes individuals with depressive symptoms and anti-depressive medication intake [69], whereas drugs acting on the dopaminergic system were excluded. Also instead of educational years, a more sensitive marker for intelligence might be a better choice for accounting for general confounding effects on working memory capacity in our sample.

Overall, our results may indeed reflect a general tendency for aspects of better working memory abilities in the good responder group as has previously been shown by other postoperative studies. Alterations in several neurocognitive domains have consistently been reported after bariatric surgery, including improvement of working memory. In a prospective study before and after bariatric surgery (RYGB (n=49) or gastric banding (n=1)), patients revealed improvements in executive functions at 3 months and 3 years after surgery [29]. Several other studies have investigated the overall improvement of various cognitive domains after bariatric surgery with mixed results. Findings for executive functions and memory were most consistent (see reviews by [27,28]), while results in most other cognitive domains were heterogeneous and depended on the study design with follow-up examination time ranging from 3 to 36 months after surgery. Moreover, ameliorated executive function, including attention, has been shown shortly i.e., 2 weeks after surgery for RYGB only but not SG, also being sustained at 3 months after surgery [70]. But not all studies confirmed an improvement in cognitive abilities after surgery. In particular, in a study comparing executive function of patients after surgery with that of waiting-list controls, null results were reported [71]. In our study, while GR showed more substantial weight loss, the postoperative group comparison compares groups with different BMI ranges (PR: 40-62 kg/m^2^, class 3 obesity; GR: 25-38 kg/m^2^, normal-weight to class 2 obesity). Therefore, we cannot exclude a general effect of elevated BMI on executive function, as was shown previously [25].

### 4.4 Postprandial secretion of gut peptides as a mediator between working memory and surgery outcome

Linking the gut-brain axis via postprandial secretion of gut peptide and working memory function is a new and explorative approach. We could show that ghrelin suppression after meal challenge mediated differences in working memory capacity, measured as IS score, by weight group after bariatric surgery. This effect was limited only to ghrelin, but not found for GLP-1 or PYY.

A direct influence of gut hormones on cognitive functions might derive from overlaps in the underlying neurobiological regulatory systems. Gut hormones e.g., ghrelin inform the brain about peripheral energy status. In addition to the hypothalamus regulating energy homeostasis, such hormones also influence eating behavior by regulating reward, motivation, and learning behavior via activation of dopaminergic midbrain pathways. These pathways, originating in the mesencephalon, project to the striatum / nucleus accumbens and prefrontal cortex and exert a direct influence on general cognitive functions such as executive functions through the release of dopamine at central neural pathway loops between the frontal brain and striatum [30], see **Figure 6**. Different cognitive functions have been associated with specific dopamine receptors [72] and dopamine levels in the mesolimbic system [73]. Further, an inverted U–shaped dynamic of dopamine signaling on working memory has been suggested, proposing an optimal dopamine level for highest working memory performance [74]. Whereas pathological eating behavior and obesity have been associated with an aberrant dopaminergic function and excessive reward-driven eating habit [75], bariatric surgery may normalize dopaminergic transmission and therefore contribute to sustained weight loss through healthier food intake behavior [10]. The importance of ghrelin and other gut hormones on postoperatively altered dopaminergic signaling has been hypothesized [76] mainly based on animal studies. Indeed, in a recent study, obese mice showed markedly altered expression of dopamine receptors after gastric bypass, compared to sham- or non-operated animals [77]. A translation of these findings to humans has produced conflicting results [78–83], which might be due to non-responding confounders (in terms of weight loss and neural adaptation) during early postoperative examinations when the steady state has not yet been established. Importantly, our findings suggest that in a stabilized weight loss maintenance state, higher performance in dopamine-dependent cognitive functions in good responders to surgery, might be associated with an altered dopaminergic state, which might be mediated by a higher response of ghrelin to food intake. In line, increased fasted ghrelin (and decreased leptin) levels 12 months after bariatric surgery (mostly RYGB) predicted better postoperative cognitive outcome [84]. Additionally, reduced plasma ghrelin levels correlated with decreased response to food cues in prefrontal brain areas using task-based functional magnetic resonance imaging (fMRI) one month after SG [85]. Interestingly, another study found no change in ghrelin levels, despite altered food cue reactivity after RYGB, yet in an early postoperative state (9 weeks after RYGB) [86]. Importantly, a limitation of all mentioned studies is that only fasting levels of gut peptides were measured, which fail to capture the postprandial dynamics of secretion. Furthermore, beyond its role in homeostatic regulation [87] ghrelin may link to cognitive function via its neuroprotective role shown in rodents [88] and humans [89]. In contrast lower ghrelin levels have been associated with aging and Alzheimer’s disease (reviewed in [90]), yet causal evidence remains scarce. Further evidence for gut hormonal interactions with cognition after bariatric surgery, including GLP-1 and PYY, remain to be investigated.

**Figure 6:**
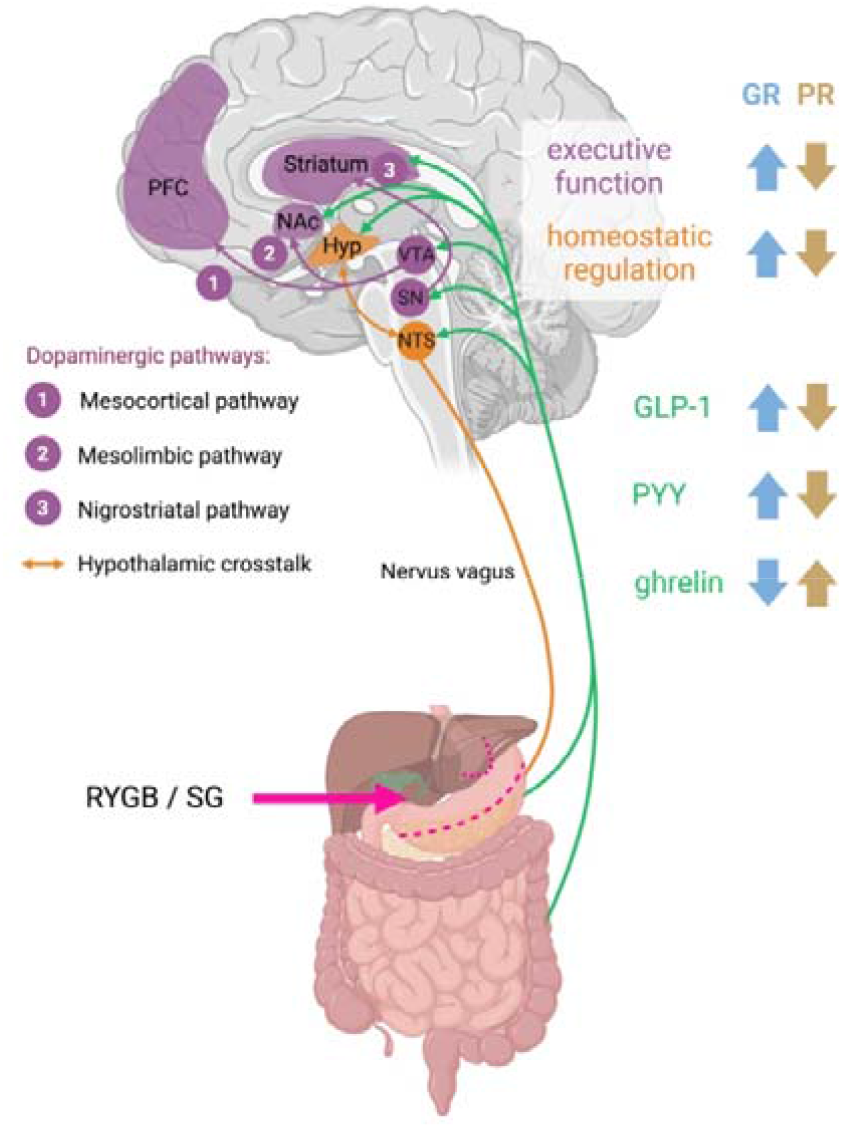
Overview of postoperative differences between good and poor responders to bariatric surgery as presented in this study. Dopamine-dependent brain pathways (in purple) involved in executive function: (1) the mesocortical pathway, (2) the mesolimbic pathway, (3) the nigrostriatal pathway. Homeostatic regulation pathways (in orange) depicting hypothalamic crosstalk between gut-brain via the vagus nerve. Gut peptide release pathways (in green) shown for GLP-1, PYY and ghrelin. Surgery-induced (pink) weight loss group differences shown as arrows for GR (blue) and PR (brown). EWL: Excessive weight loss, Hyp: hypothalamus, GR: good responder to weight loss intervention, NA: nucleus accumbens, NTS: nucleus tractus solitarii, PFC: prefrontal cortex, PR: poor responder to weight loss intervention, RYGB: Roux-en-Y gastric bypass, SG: sleeve gastrectomy, SN: substantia nigra, VTA: ventral tegmental area. Created in Biorender.com.

Notably, ghrelin mediated only IS score, but not MIS score. This might be due to the inclusion of possibly confounding, not working memory-specific distractor unit inherent to MIS score calculation, especially in the light of a low-power statistical approach and rather small effect sizes.

Taken together, our results suggest that altered secretion patterns of gut peptides are associated with weight loss after bariatric surgery, which may relate to altered neuronal pathways involved in homeostatic, hedonic and cognitive control. Although these findings must be interpreted with caution given the exploratory cross-sectional approach, a mechanistic link between successful weight loss, strong gut-hormonal response and distinct dopamine-controlled eating behavior can be assumed and awaits further, more comprehensive investigation.

### 4.5 High variability in treatment success and preoperative predictors for treatment success

While the underlying mechanisms of the weight loss effect of bariatric surgery have been steadily investigated over the past 10 years, the causes of high interindividual variability in the outcome of the surgical intervention remain insufficiently understood, leaving 20% of patients without long-term treatment success. Prediction analyses of therapy success at 12 months in more than 73.000 patients found surgery type (44.8%) and preoperative BMI (18.5%) to be most influential, while age and diabetes were rather non-influential (0.8 and 0.4%, respectively) and 34.2% of the variance was left unexplained [91]. Smaller studies also identified preoperative characteristics such as younger age and lower BMI [92,93], but also factors like healthier eating behavior, lower dietary restraint and disinhibition [94] to be predictors for long-term treatment success of bariatric surgery. In our study we also identified preoperative group BMI differences. Importantly, the absolute change from pre- to post-surgery BMI was significantly different between both groups. Preoperative BMI differences were also shown in other extreme group studies [34], but not in all [33,55]. In addition to BMI, we found further pre- and postoperative group differences for serum markers. In particular, glucose metabolism (HbA1c, insulin, HOMA-IR) and non-specific inflammation (CRP) were significantly different before and after surgery between groups, yet surgery-induced improvements were comparable. Lipid metabolism showed less preoperative differences, but distinct group differences after surgery, for HDL and triglycerides in good responders. Pre- and postoperative parameters of weight, metabolism and inflammation might be possible confounders of our analyses, which have to be considered in future studies. Ultimately, differences in personality traits, eating scores and habits were identified at post-surgery timepoint, i.e., higher extraversion and lower eating disorder scores (all EDE-Q subscales) in good responders. Those differences could equally well have been present already at pre-surgery timepoint or only have become apparent post-surgery - leaving the question of causality open.

Overall, predicting therapy success of surgery prior to treatment is a promising goal to optimize individual treatment options and outcome. One such predictive model has been proposed in a sample of >450 patients in Brazil, relying on age, and medical information, including indications on non-alcoholic fatty liver disease and cardiovascular risk factors, and the use of antihypertensive drugs [95]. Indeed, possible mechanisms for the observed high variability in surgery outcome, in our sample ranging from 0-100% EWL, include gut hormonal signaling [96] and cognitive function, yet other important metabolic factors have been disregarded in our analysis. For instance, other gut-brain-related endocrine pathways, including bile acids and liver metabolism [97] and inflammation [98], undergo drastic post-surgery adaptations that contribute to improved nutrient processing [99]. For a subsample (n=23) of the herein presented data, primary and secondary serum bile acids showed distinct profiles in GR vs. PR at fasted and postprandial state, whereas gut hormones did not, possibly due to limited statistical power [100]. Also, recent studies have highlighted the role of the vagus nerve in mediating the effects of bariatric surgery [101,102]. In addition, improvements in gut microbial diversity and gut metabolites have been noted after RYGB (in humans [103], in humans [104]). Indeed, analysis of the gut microbiome in relation to treatment success has been performed previously in a subsample of our study: specific microbiotal genera, including *Parabacteroides* (phylum: *Bacteroidetes*), linked to unhealthy eating behavior in an BMI-matched overweight sample, and was associated with treatment success after surgery [105]. Further, a reduction of hypothalamic inflammation and leptin resistance after surgery was shown in mice, acting through gut-microglia-neuron-crosstalk [65]. Overall, bottom-up gut-brain signaling has been proposed to influence higher brain functions, including reward and emotional signaling, that may also lead to adaptations in (food-seeking) behavior after bariatric surgery [106].

In sum, our results help explaining variability in treatment success after bariatric surgery, with a focus on gut hormones and working memory capacity, yet causality remains to be determined.

### 4.6 Strengths & Limitations

This study has several strengths. Firstly, the long post-surgery interval of about 5 years on average suggests that patients were in a post-surgery steady state. This is reflected in the weight trajectories which show persistent group differences between the weight loss groups and therefore justifies the herein used statistical approach of an extreme group design. Secondly, we collected extensive and detailed biomarker data at baseline and during a meal challenge paradigm to able to precisely monitor dynamics of metabolic regulation after meal intake. Thirdly, our measures of interest were carefully selected: For investigating postprandial profile, incremental AUC analysis is favorable to baseline AUC, because of the temporal aspect of hormonal secretion and the differential metabolic effect of food intake. Ultimately, we assessed working memory function, marking a rather novel approach to investigate cognitive mechanisms related to postoperative treatment success.

Certain limitations of the study have to be mentioned. Sample size was rather limited regarding complex statistical models and correcting for multiple confounding factors, whilst expecting rather small to moderate effect sizes of group differences. Also, the main analysis was based on cross-sectional data from a post-hoc design. Therefore, we cannot draw causal conclusions about whether identified group differences in gut hormonal or cognitive function were already present before the surgery or whether bariatric surgery improved executive function. Also, individuals selected for bariatric surgery are at risk for multiple co-morbidities, medication use and depressive symptoms, which highlights difficulties in precisely matching groups or accounting for all those factors. Our sample has a high proportion of female patients, which reflects overall higher numbers of treatments in females [107]. Importantly, preoperative baseline differences in weight and metabolic health may have confounded the presented postoperative group comparisons. Further, no control group, such as very-low calorie diet or peptide infusion treatments, was included, which would have been preferable to differentiate between the effect of weight loss per se compared to bariatric surgery-induced changes. Indeed, our analysis lacks more direct neurobiological marker of cognition, e.g., dopamine receptor-imaging.

### 4.7 Outlook

In the future, imaging studies targeting reward-related brain correlates, cognitive paradigms during functional magnetic resonance imaging or with PET radiotracer imaging could reveal more direct evidence for postoperative alterations in dopamine signaling [108]. Prospectively, dopamine-related targets might be of relevance for developing pharmacological therapies in obesity treatment [109]. Further, in particular for bariatric surgery where samples are mostly of small size, data pooling and meta-analyses (e.g., meta-regression across 22 reports [110]; meta-analysis across 17 studies [111]) will help to draw more robust conclusions also on gut-brain relationships. Naturally, prospective studies with a longitudinal design and multiple follow-up datapoints with adequate control groups will help to investigate the causal link between improvements in cognitive impairment and changes in hormonal responses to food intake after bariatric surgery. Our study proposes preliminary evidence for a mechanistic route between gut hormones and cognition linked to divergent therapy success after bariatric surgery, yet further research is needed to move towards clinical relevance and individualized treatment recommendations.

## Data Availability

All data produced in the present study are available upon reasonable request to the authors.

## 5. Acknowledgement

We thank Arno Villringer and coworkers of the Department of Neurology (Max Planck Institute for Human Cognitive and Brain Sciences, Leipzig, Germany) for providing laboratory space and additional resources. We also thank Tatjana Schütz and the other staff of the Integrated Research and Treatment Center AdiposityDiseases for providing excerpts from the bariatric surgery database. We further thank Suse Prejawa and Ramona Menger for administrative support; Bettina Johst and Steven Kalinke for cognitive task programming; and Anja Moll for technical-laboratory support. Furthermore, we thank Larissa Pauli and Denise Linke for testing participants and Pauline Baßler for test data digitization. Special thanks are extended to Lieneke Janssen and Maria Pössel for help in revising, methodological and statistical work; and to the remainder O’Brain team for general support. This work was funded by the German Research Foundation (project number 209933838 – SFB 1052, subproject A5 (AH)) and the Integrated Research and Treatment Center AdiposityDiseases, Federal Ministry of Education and Research (BMBF), Germany (FKZ: 01E01501 (AH)). Open access funding enabled and organized by Projekt DEAL.

## 7. Repository / Supplemental data

### 7.1 Recruitment/Exclusion Criteria

From a database of 550 patients that received laparoscopic RYGB or laparoscopic SG at the Leipzig University Medical Centre between 2007 and 2016, we contacted patients according to their weight loss and a minimum of 24 months since surgery (n = 222), of which 115 were not interested or did not live close by, 30 could not be located and four had passed away by the time of the contact. Out of 53 invited patients, five tested participants were excluded in retrospect for not meeting exclusion criteria. Two with a history of cerebral trauma, two for showing higher weight loss than anticipated (EWL > 50 %) at test day due to further gut surgery, one with a history of medical-induced-adiposity by long-term use of cortisone and one session was cancelled right after the beginning due to health issues. Furthermore, one participant disliked the test meal leading to missing hormonal data and one participant had to leave before the cognitive assessment due to personal reasons.

**Supplementary Figure 1:**
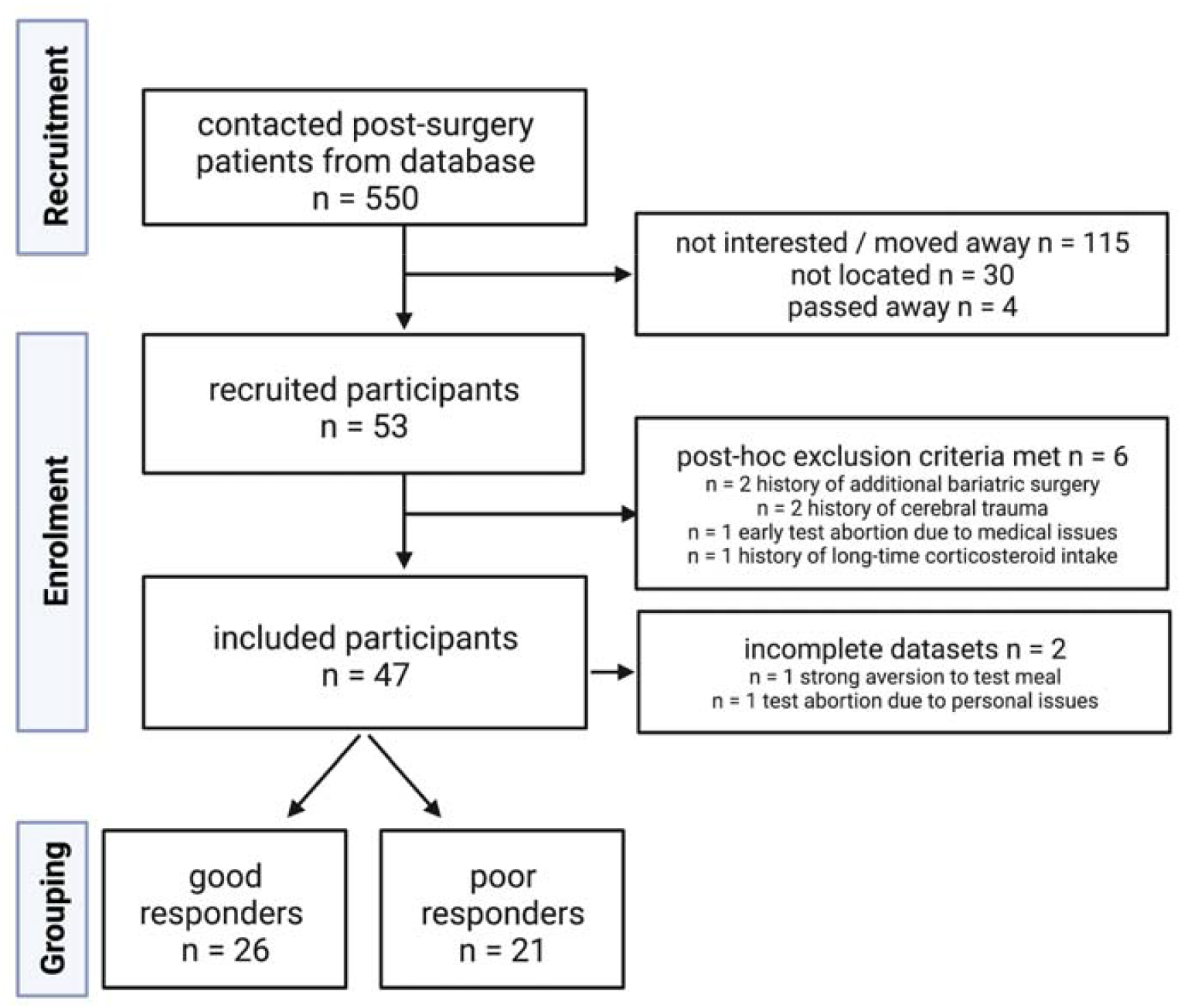
Chart of study design.

### 7.2 Surgical procedures

Before surgery, all patients underwent a hypocaloric diet, rich in protein, including commercial protein shakes, for 2 weeks. Bariatric procedures were performed standardized according to IFSO guidelines, Roux-en-Y gastric bypass with a 150cm alimentary und 50cm biliopancreatic limb, Sleeve Gastrectomy with a 34f calibration tube. The indication for surgery was given interdisciplinary following the German S3- and IFSO guideline.

### 7.3 Medication use

Within the screened population, participants without relevant comorbidities and medications represented the minority. More than half all of participants (n = 28) were taking antihypertensive medication at test date. For diabetic treatment, insulin was used by six, oral antidiabetic drugs by five and subcutaneous drugs by two participants. Frequently used medications included proton-pump inhibitors (n = 15), mild pain drugs (n = 10), thyroid hormones (n = 7), gout medication (Allopurinol, n = 7), antidepressants (n = 5), and statins (n = 4). Infrequent used medications (n ≤ 3) included neuropathic pain killers, weak and strong opioids, anticonvulsants, neuroleptics, anticoagulants, antiplatelet drugs, antihistaminergic drugs, triptan, contraceptives, antiarrhythmics, antirheumatic medication, muscle relaxants, zopiclone and ursodeoxycholic acid. Additional assessed relevant comorbidities included (n): Osteoarthritic symptoms (30), cholelithiasis (2), migraine (2), circulatory disorders (5), congestive heart failure (2), atrial fibrillation (2), autoimmune diseases (4), gynecological tumors (3) and one case of epilepsy.

### 7.4 Curation of GI peptides plasma levels

For ghrelin, invalid measures returned in 12 cases (2 × 0 / 30 min, 1 × 15min, 3 × 60 min, 4 × 120 min). Two persons were excluded from GLP-1 analyses because of the intake of GLP-1 agonists. Ceiling and floor effect occurred for GLP-1 > 2300 pg/ml in 11 cases (4 × 15 min, 7 × 30 min) and for PYY < 41 pg/ml in 3 cases (3 × 0 min). To avoid further data loss, we did include cases with ceiling effect at their original value and cases with floor effect at 50% of their value. The number of remaining data sets is stated in the **Supplementary Table 1** below.

**Supplementary Table 1:**
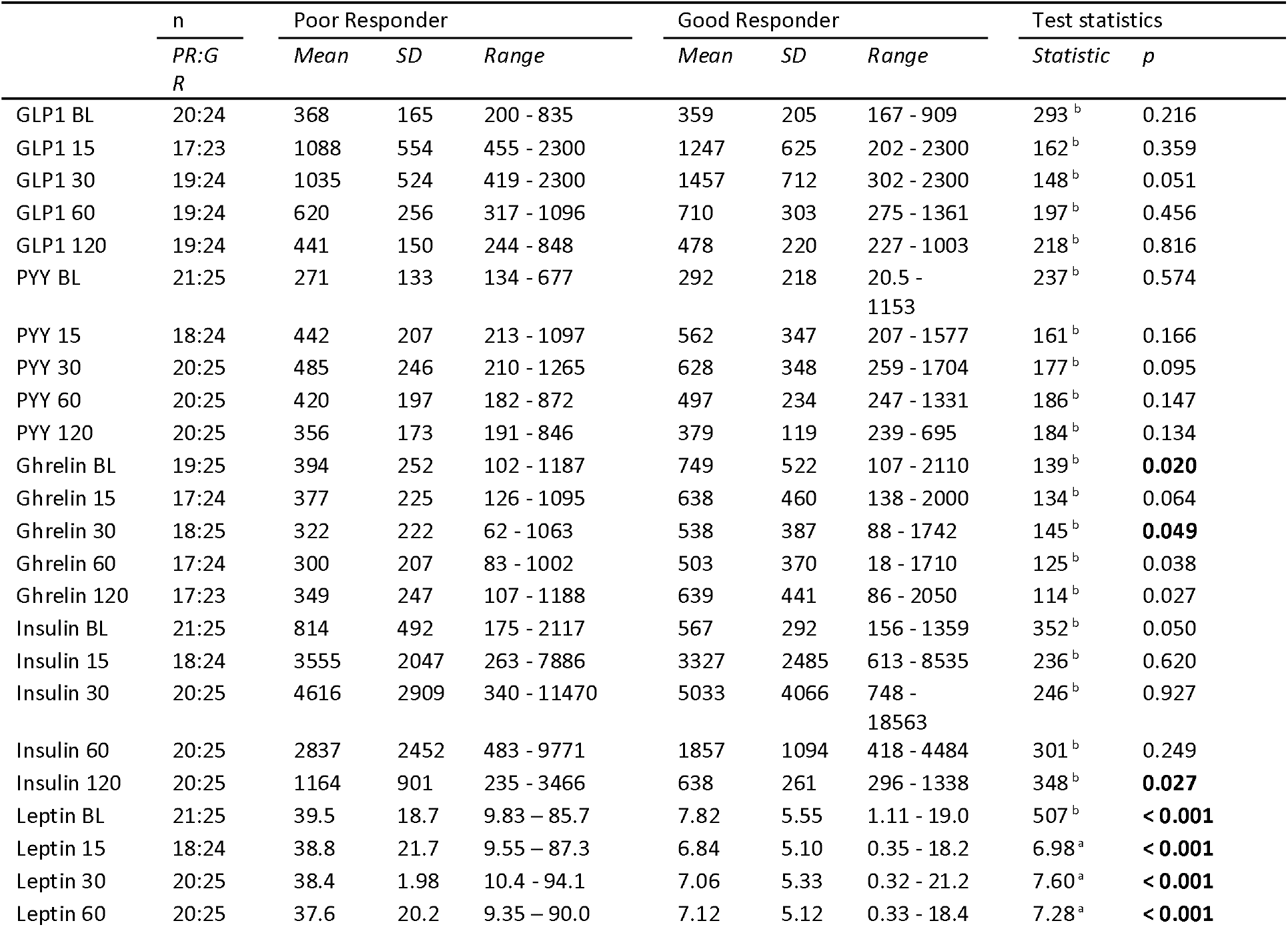
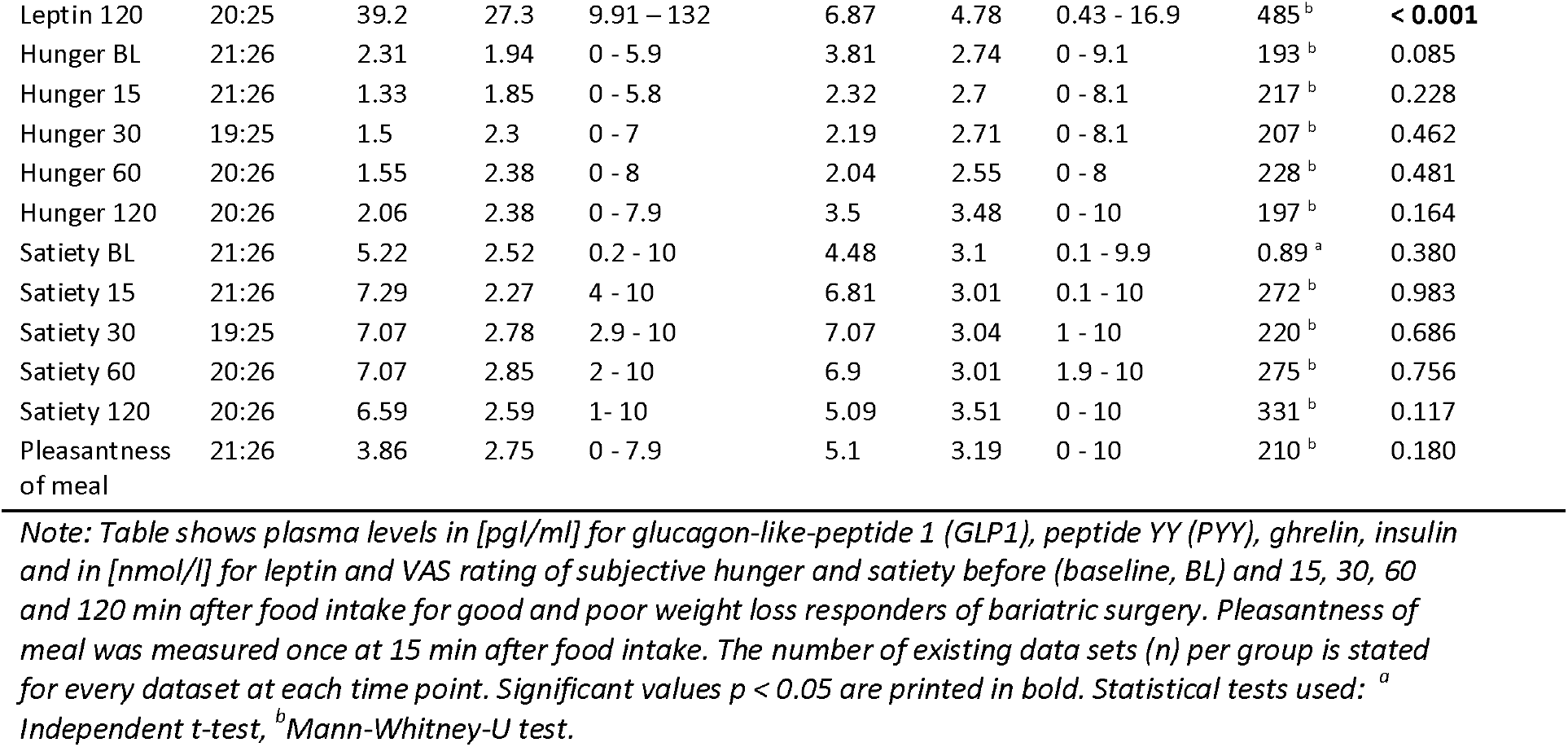
Gut hormones and VAS ratings

**Supplementary Figure 2:**
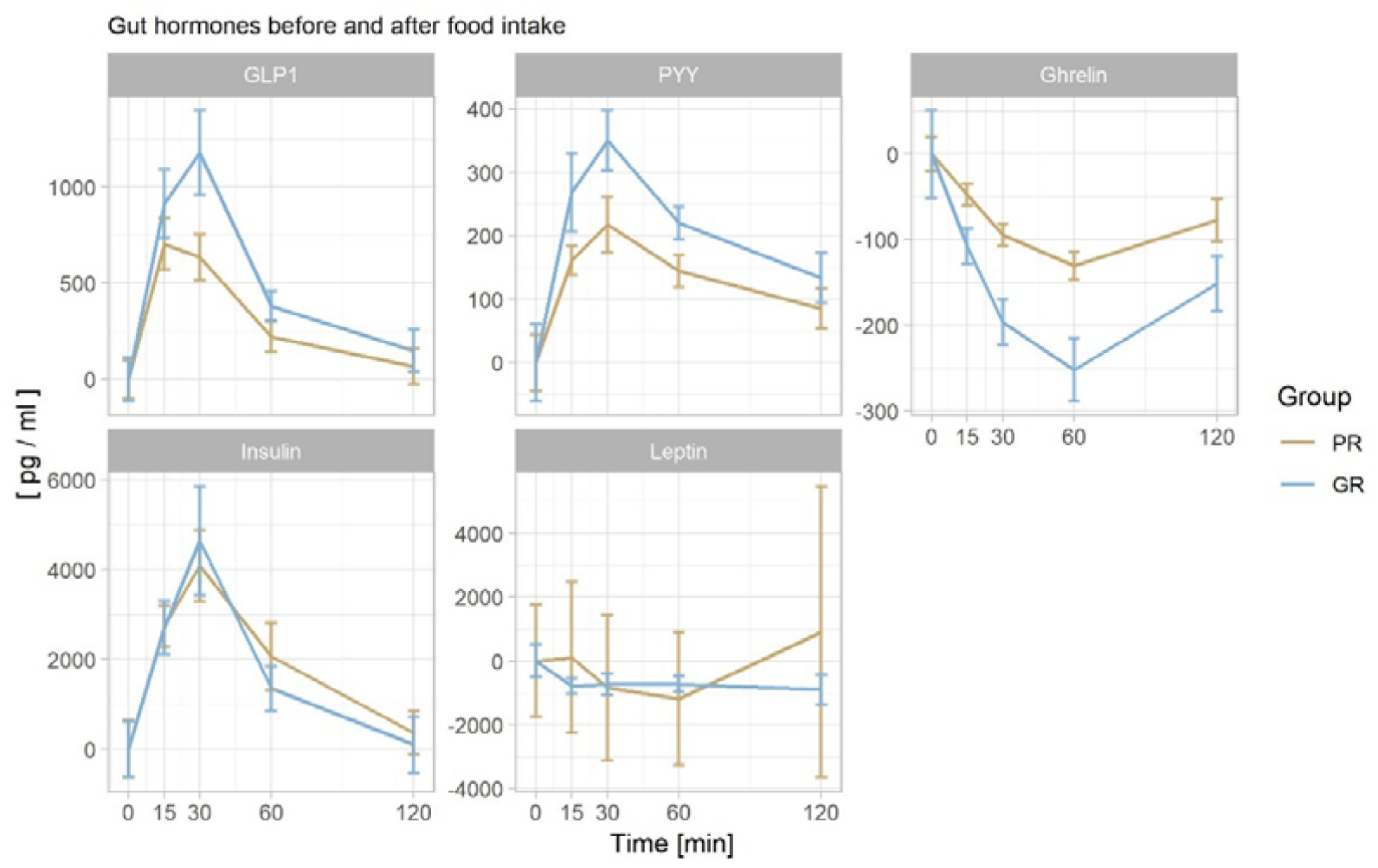
Figure representing (baseline-corrected) plasma level of glucagon-like-peptide 1 (GLP1), peptide YY (PYY), ghrelin, insulin, leptin in response to food intake at 15, 30, 60 and 120 min for good (GR) and poor weight loss responders (PR) of bariatric surgery. Error bars indicate within-subject 95% confidence interval. Data includes outliers.

**Supplementary Table 2:** Incremental Area under the curve (iAUC) for measured plasma levels of gut hormones and self-reported feelings of hunger and satiety before and after food intake

| iAUC | Poor Responder |  |  | Good Responder |  |  | Test statistics |  |
| --- | --- | --- | --- | --- | --- | --- | --- | --- |
|  | Mean | SD | Range | Mean | SD | Range | Statistic | p |
| GLP1 (17:21) | 6.25 | 3.64 | 1.08 - 13.2 | 10.5 | 5.12 | 1.33 - 20.5 | -2.86<sup>a</sup> | **0.007** |
| PYY (18:22) | 2.28 | 1.68 | 0.09 - 6.18 | 3.65 | 2.5 | 0.20 - 10.5 | 130<sup>b</sup> | 0.066 |
| Ghrelin (16:21) | -1.25 | 0.66 | -2.63 - -0.15 | -2.53 | 1.99 | -6.33 - 0.03 | 2.46 <sup>a</sup> | **0.019** |
| Insulin (18:22) | 37.5 | 25.3 | 1.76 - 105 | 37.4 | 30.3 | 4.10 - 133 | 199 <sup>b</sup> | 0.989 |
| Leptin (18:22) | -6.90 | 45.9 | -73.4 - 150 | -10.3 | 18.8 | -72.5 - 12.7 | 201 <sup>b</sup> | 0.946 |
| Hunger (19:25) | -12.5 | 32.8 | -66.1 - 66.9 | -18.1 | 28.8 | -83.3 - 21.9 | 243 <sup>b</sup> | 0.906 |
| Satiety (19:25) | 30.6 | 37.7 | -10.7 - 132 | 26.6 | 34.4 | -52.0 - 105 | 242 <sup>b</sup> | 0.924 |

Note: Incremental area under the curve (iAUC) indicates gut-hormonal response starting from fasted value (0 min) and is expressed in [ $\mu\text{g} / \text{ml} \times \text{min}$ ]. Number of compared datasets are stated in brackets (PR:GR) for each parameter. Significant values $p < 0.05$ are printed in bold. Statistical tests used: <sup>a</sup> Independent t-test, <sup>b</sup> Mann-Whitney-U test.

**Supplementary Table 3:** Linear Mixed Effect (LME) models for plasma hormone level (GLP-1, PYY, Ghrelin, Leptin and Insulin) and VAS scores (hunger and satiety) before and after test food intake.

|  | Time |  | Group |  | Time x Group |  |
| --- | --- | --- | --- | --- | --- | --- |
| | $\chi^2$ | $p$ | $\chi^2$ | $p$ | $\chi^2$ | $p$ |
| GLP-1 | 239.40 | **< 0.001** | 0.71 | 0.399 | 10.72 | **0.030** |
| PYY | 129.49 | **< 0.001** | 1.57 | 0.210 | 11.10 | **0.025** |
| Ghrelin | 115.47 | **< 0.001** | 6.15 | **0.013** | 17.72 | **0.001** |
| Leptin | 17.84 | **0.001** | 42.13 | **< 0.001** | 6.09 | 0.192 |
| Insulin | 236.44 | **< 0.001** | 1.83 | 0.177 | 5.39 | 0.250 |
| Hunger | 25.42 | **< 0.001** | 2.80 | 0.095 | 3.41 | 0.492 |
| Satiety | 51.80 | **< 0.001** | 0.70 | 0.403 | 4.93 | 0.295 |

Note: $\chi^2$ and $p$ -values are reported for main effects (Time and Group) and interaction effect (Time x Group) in comparison to the same model without the interaction term, using maximum likelihood estimation, with 4, 1 and 4 degrees of freedom, respectively. To fit test assumptions, log-transformation was performed for GLP1, insulin and leptin, as well as influential case exclusion ( $n=1$ each) for insulin and leptin. Significant values $p < 0.05$ are printed in bold.

**Supplementary Figure 3:**
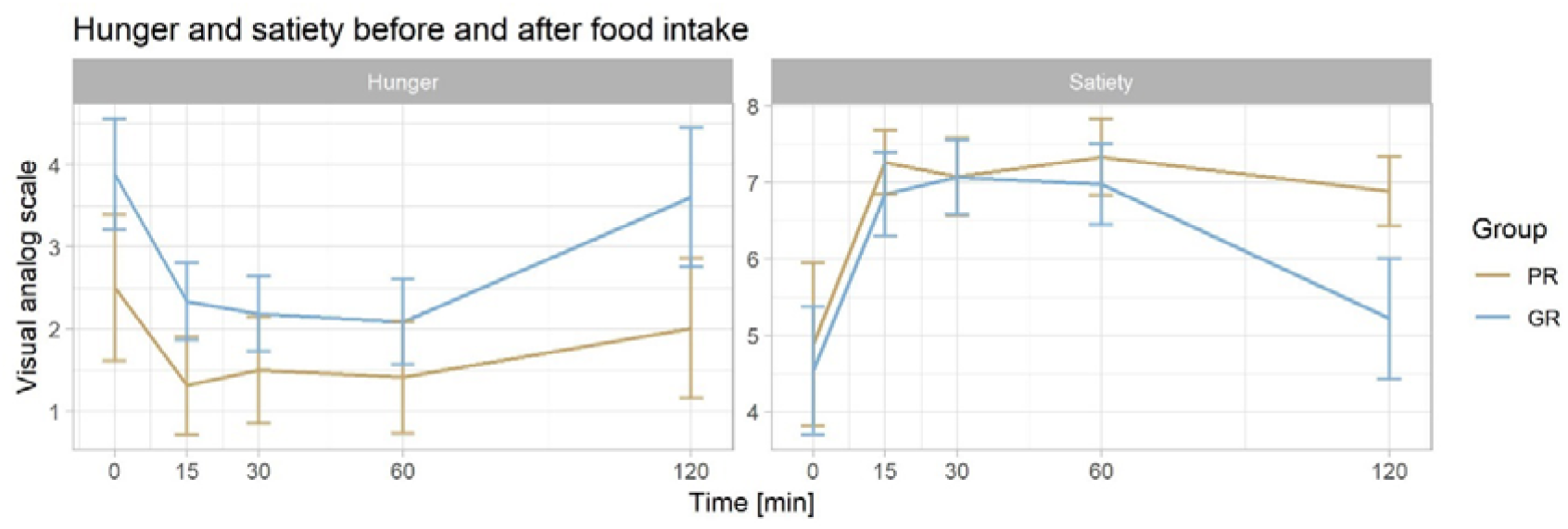
Self-reported feelings of hunger and satiety on visual analog scales (0-10) before (0 min) and 15, 30, 60 and 120 min after food intake for good (GR) and poor weight loss responders (PR) of bariatric surgery. Error bars indicate within-subject 95% confidence interval.

**Supplementary Table 4:** Correlation of self-reported feelings of hunger / satiety and plasma hormone level as baseline-corrected postprandial dynamic (iAUC).

| iAUC | GLP-1 | PYY | Ghrelin | Insulin | Leptin |
| --- | --- | --- | --- | --- | --- |
| Hunger | **-0.360 \***<br>*0.028 (0.275)* | -0.23<br>*0.154 (1.0)* | **0.33 \***<br>*0.048 (0.434)* | -0.170<br>*0.281 (1.0)* | 0.2<br>*0.223 (1.0)* |
| Satiety | 0.290<br>*0.072 (0.580)* | 0.18<br>*0.276 (1.0)* | -0.12<br>*0.485 (1.0)* | 0.180<br>*0.272 (1.0)* | -0.15<br>*0.364 (1.0)* |

Note: We report Spearman's rho and $p$ value in cursive and adjusted alpha level with Holm-Bonferroni in brackets.

### 7.5 Self-reported questionnaires

Participants completed the Beck Depression Inventory (BDI-II) [112] to assess depressive symptoms, the Trier Inventory for Chronic Stress (TICS) [113], and the Fagerstroem Test for Nicotine dependency (FTND) [114] for history of smoking. Eating behavior was assessed by the Eating Disorder examination questionnaire (EDE-Q; quantifies restraint, eating concern, weight concern and shape Concern) [115], Dutch Eating Behavior Questionnaire (DEB-Q; quantifies restrained, emotional, and external eating) [116], and Dietary Fat and Free Sugar Questionnaire (DFS-Q; quantifies fat and sugar intake) [117]. The personality inventory NEO-FFI [118] assessed the five personality traits openness to experience, conscientiousness, extraversion, agreeableness, and neuroticism. The Difficulties in Emotion Regulation Scale (DERS) [119,120] measured emotional regulation, the Urgency, Premeditation, Perseverance, Sensation Seeking Impulse Behavior Scale (UPPS) measured impulsivity [121,122], the Monetary Choice Questionnaire (MCQ) [123,124] measured delay gratification, and the Behavioral Inhibition and Behavioral Activation System Scales measured the motivation to avoid negative or approach positive stimuli (BIS/BAS) [125,126]. All questionnaires except for the BDI, which was administered on paper, were administered using the online survey tool LimeSurvey (LimeSurvey GmbH, Hamburg, Germany). Scores were computed according to the respective manuals using ‘memisc’ package [127]. Also we assessed years of education using a sum score of approximated numbers of years for each type of scholar and professional graduation in Germany.

**Supplementary Table 5:** Psychosocial characteristics of good and poor responder of bariatric surgery.

|  | Poor Responder (21) |  |  | Good Responder (26) |  |  | Test statistics |  |
| --- | --- | --- | --- | --- | --- | --- | --- | --- |
|  | Mean | SD | Range | Mean | SD | Range | Statistic | p |
| Education years | 13.5 | 1.6 | 12 - 17 | 13.7 | 1.86 | 10 - 17 | 229<sup>b</sup> | 0.280 |
| Smokers (n / %) | 4 | 19.5 |  | 6 | 23.1 |  | 0.11<sup>c</sup> | 0.737 |
| Fagerstroem (4:6) | 1.50 | 0.58 | 1 - 2 | 4 | 2 | 1 - 7 | 21.0<sup>b</sup> | 0.065 |
| BDI | 9.29 | 6.56 | 0 - 21 | 6.27 | 4.44 | 1 - 18 | 346<sup>b</sup> | 0.117 |
| DEB-Q-EE | 2.09 | 0.94 | 1 - 4.4 | 2.13 | 0.98 | 1 - 4.7 | 276<sup>b</sup> | 0.966 |
| EDE-Q | 2.47 | 0.89 | 0.78 - 4.5 | 1.21 | 0.93 | 0 - 4.04 | 78.0<sup>b</sup> | **<0.001** |
| Restraint | 1.91 | 1.62 | 0 - 6 | 0.98 | 1.31 | 0 - 4.2 | 168<sup>b</sup> | **0.022** |
| Eating concern | 0.97 | 0.96 | 0 - 3.4 | 0.48 | 0.57 | 0 - 2.2 | 176<sup>b</sup> | **0.035** |
| Weight concern | 3.11 | 1.12 | 0.8 - 5.2 | 1.52 | 1.35 | 0 - 5.2 | 97.0<sup>b</sup> | **<0.001** |
| Shape concern | 3.89 | 1.17 | 1.38 - 5.5 | 1.88 | 1.16 | 0 - 4.75 | 67.0<sup>b</sup> | **<0.001** |
| DFS-Q | 46.2 | 7.56 | 34 - 61 | 51.6 | 11.1 | 26 - 74 | 1.90<sup>a</sup> | 0.064 |
| Fat | 23.2 | 4.03 | 17 - 30 | 24.6 | 4.9 | 12 - 33 | 1.07<sup>a</sup> | 0.290 |
| Sugar | 9.71 | 2.94 | 6 - 16 | 10.5 | 3.51 | 6 - 17 | 313<sup>b</sup> | 0.395 |
| Fat & sugar | 13.3 | 3.89 | 8 - 22 | 16.5 | 4.68 | 8 - 26 | 390<sup>b</sup> | **0.013** |
| NEO-FFI | 90.2 | 6.85 | 81 - 105 | 95.7 | 8.8 | 82 - 112 | 2.31<sup>a</sup> | **0.025** |
| Extraversion | 3.02 | 0.65 | 1.67 - 4.7 | 3.44 | 0.63 | 2.8 - 4.3 | 383<sup>b</sup> | **0.019** |
| Conscientiousness | 3.71 | 0.33 | 3.0 - 4.3 | 3.87 | 0.39 | 3.0 - 4.7 | 1.47<sup>a</sup> | 0.147 |
| Neuroticism | 2.89 | 0.56 | 2 - 4.17 | 2.96 | 0.85 | 1.5 - 4.8 | 0.34<sup>a</sup> | 0.737 |
| Openness | 2.93 | 0.55 | 1.5 - 3.8 | 3.15 | 0.43 | 2.5 - 4.3 | 329<sup>b</sup> | 0.233 |
| Agreeableness | 2.49 | 0.5 | 1.5 - 3.5 | 2.52 | 0.6 | 1.3 - 3.7 | 0.17<sup>a</sup> | 0.869 |
| TICS | 14.8 | 8.38 | 0 - 35 | 17.1 | 7.74 | 7 - 35 | 317<sup>b</sup> | 0.356 |
| DERS | 75.1 | 13.9 | 56 - 104 | 74.1 | 15.8 | 41 - 102 | -0.21<sup>a</sup> | 0.833 |
| Awareness | 19.14 | 5.42 | 9 - 30 | 17.42 | 4.92 | 6 - 27 | -1.14<sup>a</sup> | 0.261 |
| Clarity | 9.29 | 3.12 | 5 - 16 | 9.65 | 3.11 | 5 - 17 | 0.40<sup>a</sup> | 0.689 |
| Goals | 10.76 | 3.55 | 5 - 16 | 9.69 | 3.62 | 05 - 18 | -1.02<sup>a</sup> | 0.315 |
| Impulsivity | 10.29 | 2.51 | 6 - 16 | 10.19 | 3.21 | 06 - 16 | 268<sup>b</sup> | 0.923 |
| Non-Acceptance | 11.57 | 5.19 | 6 - 24 | 12.46 | 3.72 | 7 - 20 | 327<sup>b</sup> | 0.250 |
| Strategies | 14 | 4.67 | 8 - 27 | 14.69 | 5.18 | 8 -24 | 290<sup>b</sup> | 0.723 |
| UPPS | 92.3 | 7.9 | 78 - 108 | 94.5 | 13.3 | 74 - 129 | 0.65<sup>a</sup> | 0.521 |
| BIS-BAS | 70.5 | 8.1 | 52 - 83 | 71.2 | 5.27 | 62 - 85 | 0.37<sup>a</sup> | 0.716 |
| MCQ | 0.06 | 0.08 | 0 - 0.25 | 0.05 | 0.08 | 0 - 0.25 | 273<sup>b</sup> | 1.000 |

Note: Smoking behavior (Fagerstroem), depression (BDI), eating behavior (EDE-Q, DEB-Q-EE, DFS-Q), personality traits (NEO-FFI), emotion regulation (DERS – subscales: Awareness (Lack of Emotional Awareness), Clarity (Lack of Emotional Clarity), Goals (Difficulties engaging in goal-directed behavior), Impulsivity (Impulse Control Difficulties), Non-Acceptance (Non-Acceptance of emotional responses) and Strategies (Limited Access to Emotion Regulation Strategies)), chronic stress (TICS – screening scale), impulsivity (UPPS), approach-avoidance behavior (BIS-BAS) and delay of gratification (MCQ). Significant values $p < 0.05$ are printed in bold. <sup>a</sup> Independent t-test, <sup>b</sup> Mann-Whitney-U test, <sup>c</sup> Pearson's Chi<sup>2</sup>-test

### 7.6 Metabolic profile

**Supplementary Table 6:** Pre- and postoperative metabolic characteristics for good and poor weight loss responders of bariatric surgery.

|  | Poor Responder (21) |  |  | Good Responder (26) |  |  | Test statistics |  |
| --- | --- | --- | --- | --- | --- | --- | --- | --- |
|  | Mean | SD | Range | Mean | SD | Range | Statistic | p |
| *At test date* |  |  |  |  |  |  |  |  |
| HbA1c (%) | 5.97 | 0.92 | 5.0 – 8.0 | 5.14 | 0.65 | 4.4 - 7.8 | 465<sup>b</sup> | **< 0.001** |
| Fasting glucose (mmol/l) | 6.52 | 1.66 | 4.8 - 11.5 | 5.17 | 0.56 | 4.3 - 6.8 | 443<sup>b</sup> | **< 0.001** |
| Insulin (pmol/l) | 82.1 | 45.0 | 26 - 234 | 46.7 | 32.7 | 11 - 167 | 442<sup>b</sup> | **< 0.001** |
| HOMA-IR | 4.02 | 2.89 | 1.0 – 15.0 | 1.82 | 1.45 | 0.6 - 7.3 | 471<sup>b</sup> | **< 0.001** |
| LDL (mmol/l) | 2.49 | 0.47 | 1.7 - 3.3 | 2.37 | 0.56 | 1.6 – 4.0 | 327<sup>b</sup> | 0.257 |
| HDL (mmol/l) | 1.44 | 0.4 | 0.5 - 2.1 | 1.84 | 0.64 | 1.2 - 3.3 | 192<sup>b</sup> | 0.083 |
| Triglycerides (mmol/l) | 1.40 | 0.4 | 0.7 - 2.2 | 0.89 | 0.39 | 0.5 - 2.1 | 466<sup>b</sup> | **< 0.001** |
| Cholesterol (mmol/l) | 4.18 | 0.64 | 3.0 - 5.2 | 4.31 | 0.9 | 2.9 - 6.2 | -0.55<sup>a</sup> | 0.584 |
| ALAT (μkat/l) | 0.37 | 0.15 | 0.2 - 0.7 | 0.41 | 0.18 | 0.1 - 1 | 244<sup>b</sup> | 0.542 |
| ASAT (μkat/l) | 0.40 | 0.11 | 0.2 - 0.7 | 0.5 | 0.22 | 0.3 - 1.5 | 183<sup>b</sup> | 0.054 |
| CRP (mg/l) | 12.8 | 32.7 | 0.7 - 153 | 0.65 | 0.75 | 0.0 - 3.6 | 509<sup>b</sup> | **< 0.001** |
| IL-6 (pg/ml) (21:25) | 6.84 | 9.97 | 1.3 - 49.3 | 2.17 | 2.96 | 1.3 - 15.5 | 469<sup>b</sup> | **< 0.001** |
| *Before surgery* |  |  |  |  |  |  |  |  |
| HbA1c (17:22) | 6.44 | 1.86 | 4.9 - 11.5 | 5.56 | 0.89 | 4.2 - 7.7 | 252<sup>b</sup> | 0.068 |
| Fasting glucose (19:25) | 8.08 | 4.57 | 4.9 - 18.3 | 5.74 | 1.68 | 2.2 - 11.9 | 333<sup>b</sup> | **0.024** |
| Insulin (21:24) | 167 | 130 | 40.3 - 644 | 122 | 116 | 9.2 - 482 | 340<sup>b</sup> | **0.047** |
| HOMA-IR (19:23) | 11.2 | 18.9 | 2.5 - 87.6 | 6.27 | 8.35 | 0.3 - 38.1 | 323<sup>b</sup> | **0.009** |
| LDL (18:21) | 2.86 | 1.02 | 1.5 - 4.6 | 2.87 | 0.86 | 1.7 - 5.1 | -0.02<sup>a</sup> | 0.984 |
| HDL (18:21) | 1.09 | 0.29 | 0.5 - 1.7 | 1.18 | 0.45 | 0.6 - 2.5 | 183<sup>b</sup> | 0.877 |
| Triglycerides (18:21) | 1.88 | 1.14 | 0.9 – 5.0 | 1.29 | 0.54 | 0.5 - 3.1 | 253<sup>b</sup> | 0.074 |
| Cholesterol (18:21) | 4.74 | 1.16 | 2.7 - 7.1 | 4.55 | 1.08 | 3.0 - 7.2 | 0.51<sup>a</sup> | 0.611 |
| ALAT (21:26) | 0.69 | 0.61 | 0.2 - 3.1 | 0.65 | 0.57 | 0.2 - 3.0 | 297<sup>b</sup> | 0.615 |
| ASAT (21:26) | 0.58 | 0.37 | 0.3 - 2.0 | 0.54 | 0.26 | 0.3 - 1.4 | 299<sup>b</sup> | 0.585 |
| CRP (21:26) | 11.4 | 7.45 | 1.1 - 26.7 | 7.78 | 7.58 | 0.6 - 31.1 | 361<sup>b</sup> | 0.061 |

Note: Numbers of compared datasets in brackets (PR:GR), if incomplete.
Significant values $p < 0.05$ are printed in bold. Statistical tests used: <sup>a</sup> Independent t-test, <sup>b</sup> Mann-Whitney-U test

**Supplementary Figure 4:**
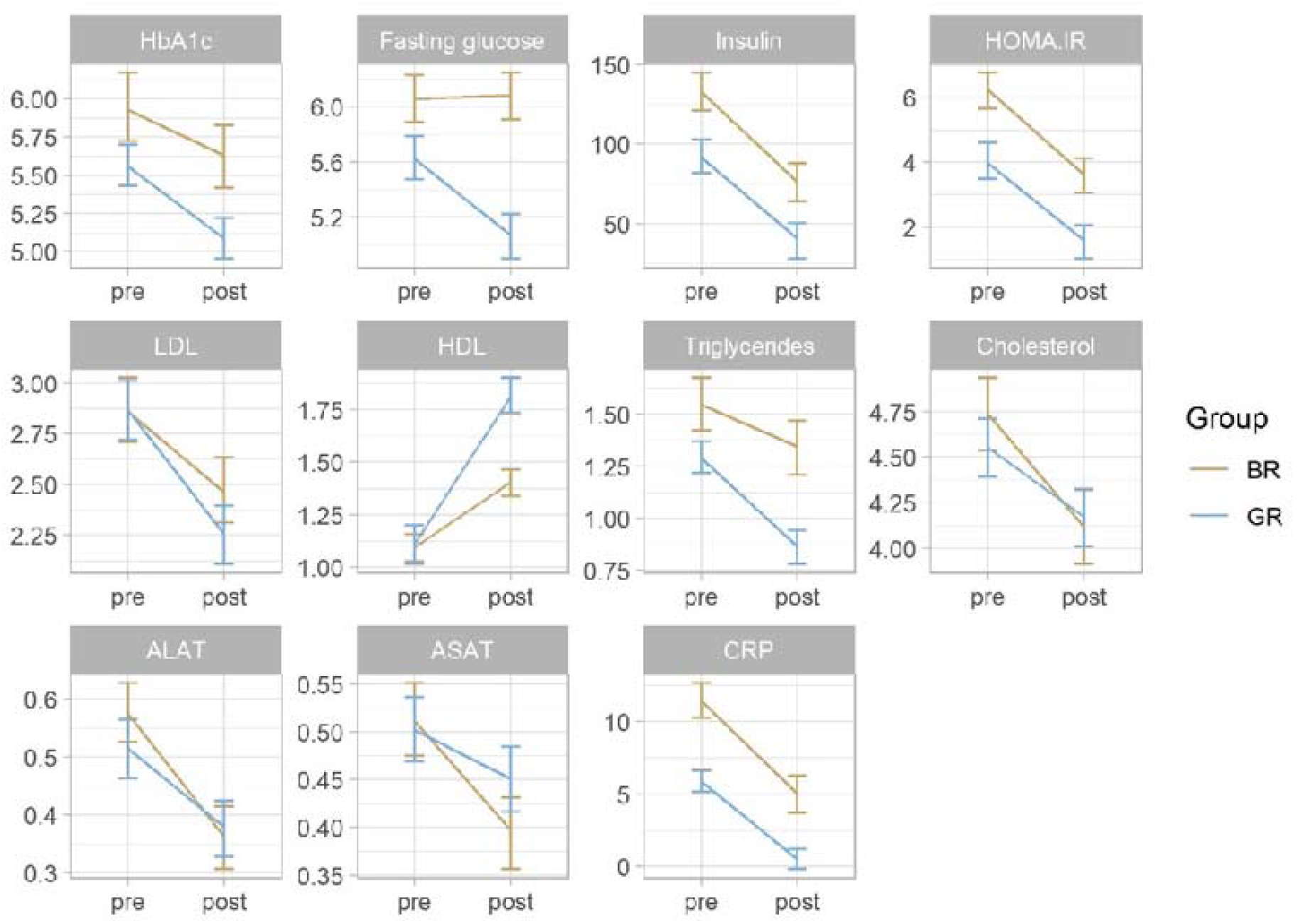
Metabolic improvement since bariatric surgery. Error bars indicate within-subject 95% confidence interval. Statistical outlier were excluded.

**Supplementary Table 7:** Linear Mixed Effect (LME) models of blood parameters over time (before to after surgery)

|  | *n* (PR:GR) |  | Time |  | Group |  | Time x Group |  |
| --- | --- | --- | --- | --- | --- | --- | --- | --- |
| | *pre* | *post* | $\chi^2$ | *p* | $\chi^2$ | *p* | $\chi^2$ | *p* |
| HbA1c | 15:22 | 20:26 | 7.00 | **0.008** | 6.90 | **0.009** | 1.00 | 0.316 |
| Fasting glucose | 16:23 | 19:26 | 4.42 | **0.035** | 8.65 | **0.003** | 4.64 | **0.031** |
| Insulin | 20:22 | 20:26 | 29.7 | **< 0.001** | 9.30 | **0.002** | 1.03 | 0.311 |
| HOMA-IR | 17:21 | 20:26 | 28.4 | **< 0.001** | 13.2 | **< 0.001** | 0.49 | 0.486 |
| LDL | 18:21 | 21:25 | 16.1 | **< 0.001** | 0.28 | 0.6 | 0.86 | 0.354 |
| HDL | 18:20 | 21:26 | 41.3 | **< 0.001** | 3.51 | **0.061** | 10.1 | **0.001** |
| Triglycerides | 16:21 | 21:26 | 14.1 | **< 0.001** | 9.77 | **0.002** | 2.7 | 0.1 |
| Cholesterol | 18:21 | 21:26 | 10.1 | **0.001** | 0.003 | 0.959 | 1.07 | 0.301 |
| ALAT | 20:24 | 21:25 | 16.3 | **< 0.001** | 0.36 | 0.551 | 1.11 | 0.292 |
| ASAT | 20:25 | 21:25 | 8.06 | **0.005** | 0.38 | 0.538 | 1.69 | 0.194 |
| CRP | 21:24 | 19:26 | 43.9 | **< 0.001** | 14.7 | **< 0.001** | 0.61 | 0.434 |

Note: Pre- and postoperative numbers of cases (*n*) are depicted after formal outlier exclusion (2.2 x interquartile range from upper / lower quartile). $\chi^2$ and *p*-value are reported for main effects (time and group) and interaction effect (time x group) in comparison to the same model without the interaction term, using maximum likelihood estimation. Significant values *p* < 0.05 are printed in bold

**Supplementary Table 8:**
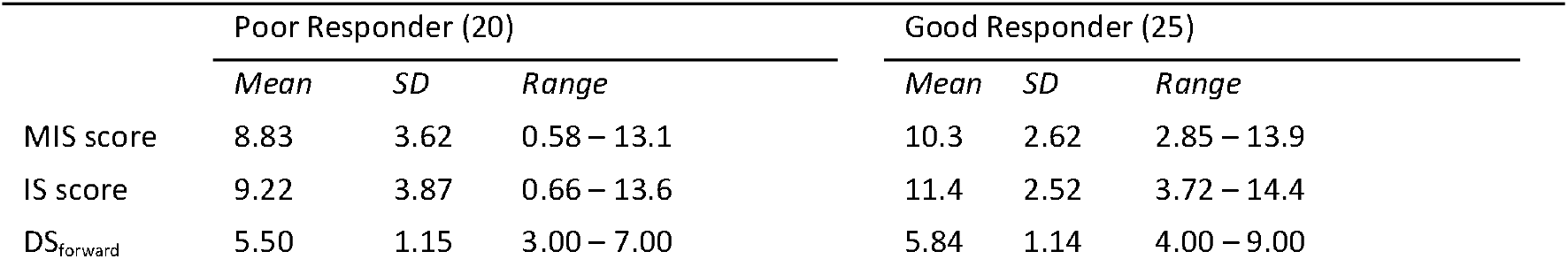
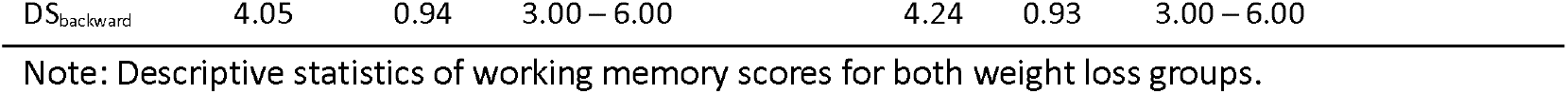
Working memory subscores.

**Supplementary Table 9:** Mediation models

| Variables | *B* | *SE* | *t* | *p* |
| --- | --- | --- | --- | --- |
| Direct effects |  |  |  |  |
| **MIS score** |  |  |  |  |
| *Total effect of predictor on outcome (path c)* |  |  |  |  |
| Weight loss → MIS score | 1.90 | 0.96 | 1.98 | 0.048 |
| *Effect of predictor on mediators (path a)* |  |  |  |  |
| Weight loss → GLP-1 | 4.16 | 1.45 | 2.87 | 0.004 |
| Weight loss → PYY | 1.14 | 0.67 | 1.70 | 0.089 |
| Weight loss → Ghrelin | -1.32 | 0.49 | -2.70 | 0.007 |
| *Effect of mediators on outcome (path b)* |  |  |  |  |
| GLP-1 → MIS score | -0.12 | 0.13 | -0.93 | 0.355 |
| PYY → MIS score | 0.62 | 0.20 | 3.08 | 0.002 |
| Ghrelin → MIS score | -0.35 | 0.25 | -1.38 | 0.167 |
| *Direct effect of predictor on outcome (path c')* |  |  |  |  |
| Weight loss (+ mediators) → MIS score | 1.21 | 1.13 | 1.07 | 0.286 |
| **IS score** |  |  |  |  |
| *Total effect of predictor on outcome (path c)* |  |  |  |  |
| Weight loss → IS score | 2.16 | 0.98 | 2.21 | 0.027 |
| *Effect of predictor on mediators (path a)* |  |  |  |  |
| Weight loss → GLP-1 | 4.19 | 1.44 | 2.91 | 0.004 |
| Weight loss → PYY | 1.14 | 0.69 | 1.66 | 0.097 |
| Weight loss → Ghrelin | -1.33 | 0.49 | -2.73 | 0.006 |
| *Effect of mediators on outcome (path b)* |  |  |  |  |
| GLP-1 → IS score | -0.15 | 0.13 | -1.22 | 0.222 |
| PYY → IS score | 0.68 | 0.22 | 3.03 | 0.002 |
| Ghrelin → IS score | -0.48 | 0.22 | -2.16 | 0.031 |
| *Direct effect of predictor on outcome (path c')* |  |  |  |  |
| Weight loss (+ mediators) → IS score | 1.47 | 1.15 | 1.28 | 0.201 |
| Indirect effects |  |  |  |  |
|  | Estimate | *SE* | LL 95% CI | UL 95% CI |
| **MIS score** |  |  |  |  |
| GLP-1 | -0.49 (-0.08) | 0.56 | -1.94 | 0.36 |
| PYY | 0.71 (0.11) | 0.46 | -0.04 | 1.78 |
| Ghrelin | 0.46 (0.07) | 0.38 | -0.07 | 1.54 |
| **IS score** |  |  |  |  |
| GLP-1 | -0.64 (-0.96) | 0.58 | -2.17 | 0.15 |
| PYY | 0.69 (0.11) | 0.46 | -0.08 | 1.78 |
| Ghrelin | **0.64 (0.10)** | **0.38** | **0.07** | **1.68** |

Note: Mediating effect of postprandial gut hormone release (GLP-1, PYY and ghrelin) on the relationship between weight loss group and working memory (MIS and IS score). Significant indirect effects (if 95% CI does not cross zero) in bold. Estimate in absolute and standardized (in brackets) values. SE: standard error; LL: lower level; UP, upper level; CI: confidence interval.

